# Within-household transmission risk of pulmonary tuberculosis in the era of universal antiretroviral therapy

**DOI:** 10.64898/2026.06.01.26354571

**Authors:** Palwasha Y Khan, Indira Govender, Nicky McCreesh, Mareca Sithole, Edwin Mkwanazi, Sedona Sweeney, Gregory Ording-Jespersen, Emily Wong, Willem Hanekom, Rein MGJ Houben, Richard G White, Theresa K Smit, Matthew J Smith, Katherine Fielding, Alison D Grant

## Abstract

**Background:** Tuberculosis remains the leading infectious cause of death worldwide. In the WHO African region, declining incidence has coincided with antiretroviral therapy (ART) scale-up, though whether this reflects reduced progression to disease or reduced transmission is unclear. We evaluated how ART and symptom status influence within-household *Mycobacterium tuberculosis complex* (MTBC) transmission risk.

**Methods:** We conducted a case-contact household study in rural South Africa, enrolling index adults with bacteriologically-confirmed pulmonary tuberculosis. MTBC immunoreactivity was measured in all child household contacts (aged 2-14 years) as a proxy measure of within-household transmission. We assessed the influence of index person ART status and symptom status and explored effect-measure modification of the association between index person HIV status and transmission risk by sex.

**Results:** Among 755 child contacts of 296 index persons, effective ART was not associated with within-household MTBC transmission risk (risk ratio [RR], 1.07; 95% CI, 0.66–1.74). Among PLHIV engaged in ART care, WHO TB four-symptom screen (WHO4SS) status was not associated with transmission risk (RR, 0.80; 95% CI, 0.43–1.47), although absence of reported cough reduced risk (RR, 0.61; 95% CI, 0.38–0.96). A pronounced interaction between sex and HIV status was observed: HIV-negative women had the highest within-household MTBC transmission risk (30.5% vs. 14.3% in women with HIV) whereas risks were similar between HIV-positive and HIV-negative men.

**Conclusions:** We found no evidence that effective ART or WHO4SS status influenced within-household MTBC transmission risk, though confidence intervals were wide. Absence of reported cough was associated with lower risk, and transmission risk was highest among child contacts of HIV-negative women. These findings suggest reported cough is a useful marker of transmission risk and that routine tuberculosis screening within ART care may reduce transmission from PLHIV; intensified efforts are nonetheless needed to achieve earlier tuberculosis detection in HIV-negative individuals.

## Background

Tuberculosis is the world’s leading cause of death from a single infectious agent.^1^ The World Health Organization (WHO) African region was the only region to exceed the End TB Strategy target of a 20% reduction in tuberculosis incidence between 2015 and 2020;^2^ a decline plausibly attributable to the scale-up of effective antiretroviral treatment (ART). Whether this reflects reduced progression from *Mycobacterium tuberculosis complex* (MTBC) infection to disease among people taking ART or decreased MTBC transmission remains unclear.^3^ In the pre- and early-ART era, tuberculosis case-contact studies consistently found that people living with HIV (PLHIV) starting tuberculosis treatment were less infectious to household contacts than their HIV-negative counterparts.^4–10^ This reduced within-household MTBC transmission risk may no longer persist, as earlier HIV diagnosis and ART initiation could alter the clinical presentation of HIV-associated tuberculosis toward a more infectious phenotype, thereby potentially increasing risk of within-household MTBC transmission.^11^ Figure 1 outlines a theoretical schema of the impact of ART on the risk of MTBC transmission.

**Figure 1.**
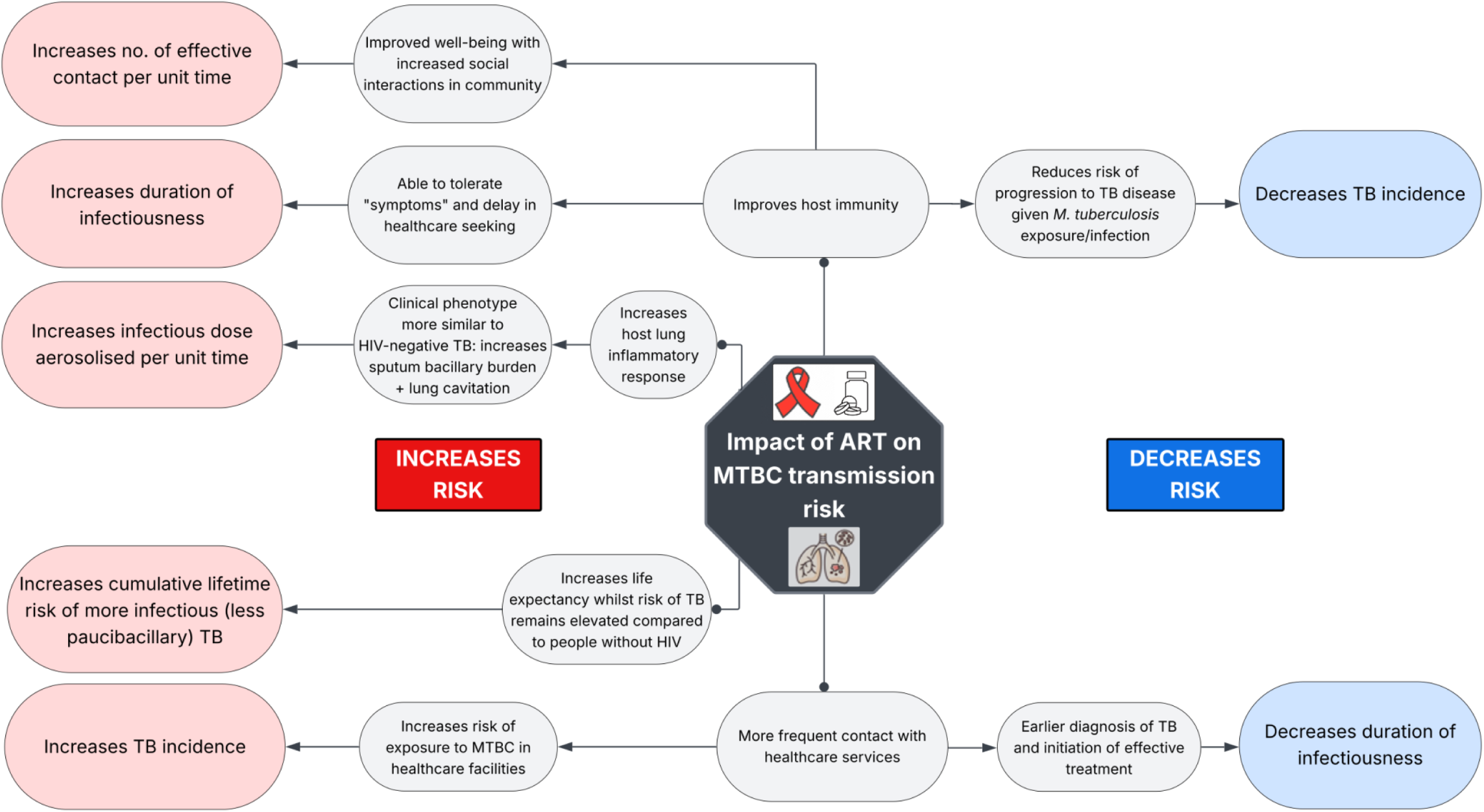
Theoretical schema of the impact of antiretroviral treatment on the risk of *Mycobacterium tuberculosis complex* (MTBC) transmission

We assessed whether index person ART status and symptom status influence within-household MTBC transmission risk to child contacts of adults with bacteriologically-confirmed pulmonary tuberculosis. Comparisons were made between (a) (a) people living with HIV (PLHIV) not on effective ART versus those on effective ART, and (b) asymptomatic versus symptomatic index persons on effective ART. We also explored (c) effect-measure modification of the association between index person HIV status and within-household transmission risk by index person sex.

## Methods

### STUDY DESIGN AND SETTING

We conducted a cross-sectional case-contact household study in the area of the Africa Health Research Institute (AHRI) health and demographic surveillance system (HDSS) in uMkhanyakude district in KwaZulu-Natal (population ∼150,000; adult HIV prevalence 33%; see map, figure S1).^12^ In a 2018 community survey, 1.4% (95% confidence interval [CI] 1.2-1.6) of adults had bacteriologically-confirmed pulmonary tuberculosis.^13^ During the study period, PLHIV on ART received routine annual, symptom-agnostic TB screening at healthcare facilities as part of programmatic care.

### STUDY PARTICIPANTS

#### Index people with pulmonary tuberculosis

We enrolled adults (≥ 15 years) with bacteriologically-confirmed pulmonary tuberculosis identified by routine services initiating tuberculosis treatment at AHRI HDSS healthcare facilities (February 2022 – December 2023). We excluded individuals with a recent history of tuberculosis treatment (within two years), those living with a household member treated for tuberculosis (within two years), and households without co-resident children aged 2 to 14 years. At enrolment, we collected demographic and clinical data, including diagnostic (programmatic) sputum Xpert MTB/RIF Ultra result, HIV and ART history, symptoms, and anthropometrics, performed digital chest radiography with computer-aided detection (CAD4TB, Delft), obtained two sputum samples for Xpert MTB/RIF Ultra and liquid culture (MGIT, Becton Dickinson), and collected blood for HIV viral load and CD4 count (in PLHIV).

#### Household contacts

Household visits were conducted approximately 6 weeks after treatment initiation. The head of household (or a proxy) completed a questionnaire on household characteristics. Children aged 2–14 years who had spent at least 2 weeks co-resident with the index adult prior to treatment initiation were eligible for QuantiFERON-TB Gold Plus (QFT) testing. A questionnaire captured demographics, symptoms, and contact intensity with the index person. Children with unknown HIV status were offered testing with an enzyme-linked immunosorbent assay. Those testing positive were linked to ART care. All children under 5 years and all HIV-positive children were referred to their primary healthcare clinic for evaluation and isoniazid preventive treatment.

Children aged 5–14 years with tuberculosis symptoms or a positive QFT were referred for assessment by a study clinician. Laboratory procedures for sample collection, processing, and interpretation of QFT are provided in the Supplementary Methods.

#### Control (non-tuberculosis-affected) households

Using the AHRI HDSS as a sampling frame, we randomly selected control adults without tuberculosis, frequency-matched to index persons by age (<40 or ≥40 years), sex, and geographical area (residence within 1 km). Identical study procedures were undertaken in control households. For further details see published protocol.^14^

### OUTCOME DEFINITION

The outcome was MTBC immunoreactivity, defined as positive QFT result in child household contacts (aged 2 – 14 years), used as a proxy for within-household MTBC transmission.

### EXPOSURES OF INTEREST

The primary exposure was index person’s ART status at tuberculosis treatment initiation, with the baseline group defined as PLHIV on effective ART (≥6 months on ART with an HIV viral load <50 copies/mL). Secondary analyses of symptom status were limited to PLHIV engaged in ART care, including both newly initiating and established on ART, as this group undergoes TB screening irrespective of symptoms as part of routine programmatic care. Index person symptoms were defined as the presence of any item from the WHO four-symptom tuberculosis screening tool (WHO4SS), i.e., reported cough, fever, night sweats, or weight loss.^15^

As an exploratory analysis, we assessed effect-measure modification of the association between index person HIV status (HIV-negative versus PLHIV) and within-household MTBC transmission risk by index person sex.

### STATISTICAL ANALYSIS

The study had 80% power to detect a relative risk of 1.67 in within-household MTBC transmission by effective ART status. Assuming 15% QFT-positivity, an intraclass coefficient (ICC) of 0.34, and an average of 2 child contacts per index person, a minimum of 340 index participants was required.

Confounding was addressed by adjusting for index person-level and household-level covariates, selected based on a pre-specified causal diagram (see Supplementary Methods and Figure S2). We set out to estimate the causal effect of the primary and secondary exposures (ART and symptom status among PLHIV) using targeted minimum loss-based estimation (TMLE; *ltmle* package in R), accounting for household clustering and incorporated ensemble machine learning for covariate adjustment to reduce the risk of model misspecification.^16,17^ For the exploratory analysis of effect-measure modification, we assessed whether index person sex modified the association between HIV status and within-household MTBC transmission risk using *MargIntTmle* package.^18–20^

As a supplementary analysis, we used multivariable generalized estimating equations (GEE) logistic regression models (with exchangeable correlation structure and robust standard errors) ^21–23^ presented in Table S5. In sensitivity analyses, we restricted to children under 11 years to minimise outcome misclassification resulting from community MTBC exposure (Table S6). Missing covariate data were handled using the missing indicator method (Table S7).

We also estimated the household-attributable risk by comparing prevalence of MTBC immunoreactivity between index (TB-affected) and control households using the same TMLE framework (see Supplementary methods). This analysis was conducted post hoc in response to peer review.

### ETHICAL CONSIDERATIONS

Written informed consent (or witnessed verbal consent for those unable to read/write) was obtained from participants or their guardians. The study was approved by ethics committees at the University of KwaZulu-Natal (BF472/19) and the London School of Hygiene & Tropical Medicine, (17728-6), and conducted in accordance with the Declaration of Helsinki.

## Results

We enrolled 362 (92.8%) eligible index persons initiating tuberculosis treatment and conducted household visits for 310 (79.5%). The number of study participants recruited was lower than our intended sample size due to COVID-19-related delays.

Among eligible child contacts aged 2–14 years, 819 (92.4%) were enrolled; 755 (85.2%) had a valid QFT result (see Supplemental methods). Study flowcharts for index and control households are provided in Figures S3 and S4.

### PARTICIPANT CHARACTERISTICS

Index participants included in the main analyses (n=296) and those without child contact data (n=66) were broadly similar, except for a higher prevalence of cavitation (20% vs. 10%) and a lower proportion reporting no symptoms (7% vs. 14%) (Table S1). Among index participants included in the main analyses (Table 1), the median age was 36 years; 50.3% were male, 25.7% had prior tuberculosis, and 65.5% (n = 194) were living with HIV, of whom 56.7% were virally suppressed.

**Table 1.**
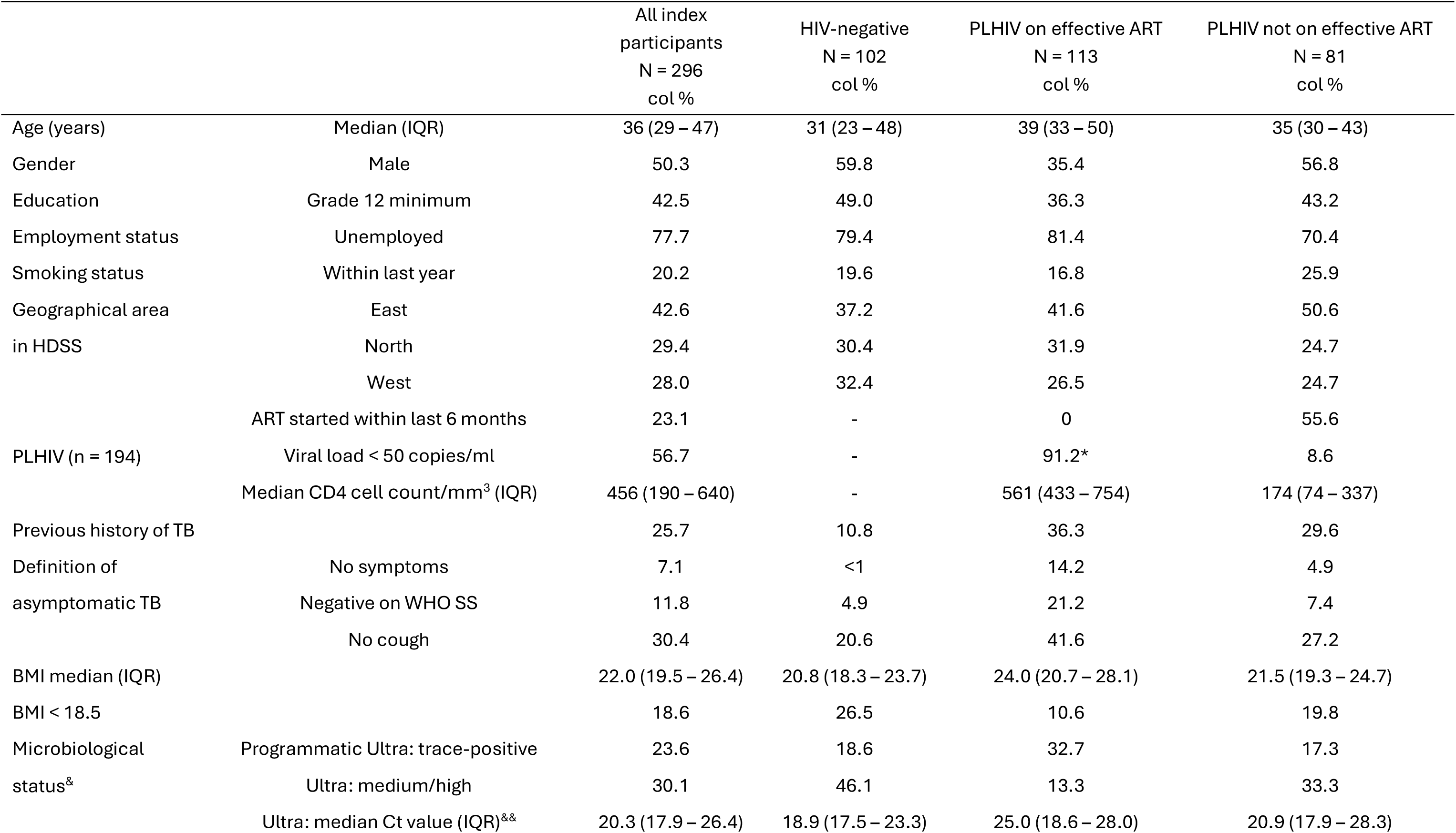
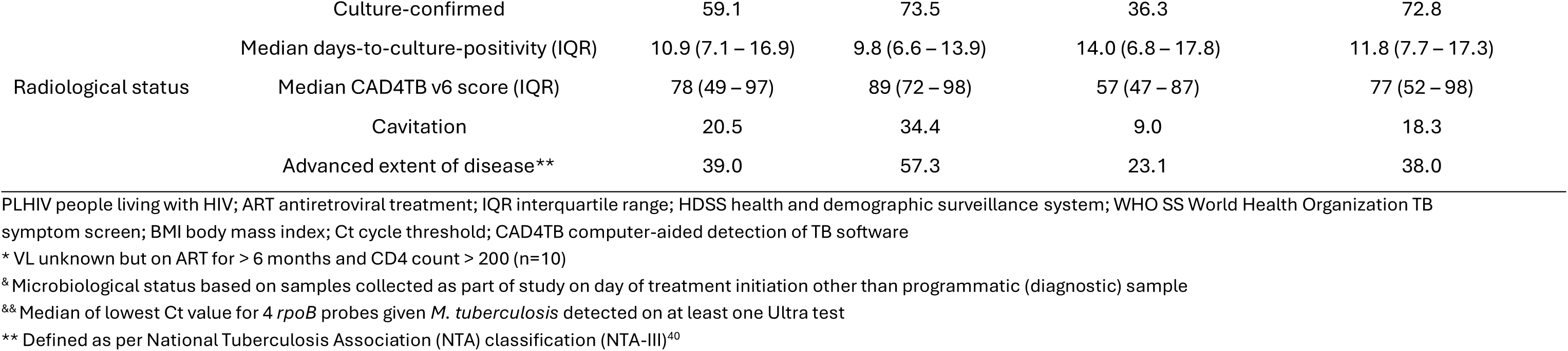
Characteristics of index participants at enrolment by HIV/ART status.

|  |  | All index<br>participants<br>N = 296<br>col % | HIV-negative<br>N = 102<br>col % | PLHIV on effective ART<br>N = 113<br>col % | PLHIV not on effective ART<br>N = 81<br>col % |
| --- | --- | --- | --- | --- | --- |
| Age (years) | Median (IQR) | 36 (29 – 47) | 31 (23 – 48) | 39 (33 – 50) | 35 (30 – 43) |
| Gender | Male | 50.3 | 59.8 | 35.4 | 56.8 |
| Education | Grade 12 minimum | 42.5 | 49.0 | 36.3 | 43.2 |
| Employment status | Unemployed | 77.7 | 79.4 | 81.4 | 70.4 |
| Smoking status | Within last year | 20.2 | 19.6 | 16.8 | 25.9 |
| Geographical area | East | 42.6 | 37.2 | 41.6 | 50.6 |
| in HDSS | North | 29.4 | 30.4 | 31.9 | 24.7 |
|  | West | 28.0 | 32.4 | 26.5 | 24.7 |
| PLHIV (n = 194) | ART started within last 6 months | 23.1 | - | 0 | 55.6 |
|  | Viral load < 50 copies/ml | 56.7 | - | 91.2* | 8.6 |
|  | Median CD4 cell count/mm<sup>3</sup> (IQR) | 456 (190 – 640) | - | 561 (433 – 754) | 174 (74 – 337) |
| Previous history of TB |  | 25.7 | 10.8 | 36.3 | 29.6 |
| Definition of | No symptoms | 7.1 | <1 | 14.2 | 4.9 |
| asymptomatic TB | Negative on WHO SS | 11.8 | 4.9 | 21.2 | 7.4 |
|  | No cough | 30.4 | 20.6 | 41.6 | 27.2 |
| BMI median (IQR) |  | 22.0 (19.5 – 26.4) | 20.8 (18.3 – 23.7) | 24.0 (20.7 – 28.1) | 21.5 (19.3 – 24.7) |
| BMI < 18.5 |  | 18.6 | 26.5 | 10.6 | 19.8 |
| Microbiological | Programmatic Ultra: trace-positive | 23.6 | 18.6 | 32.7 | 17.3 |
| status<sup>&</sup> | Ultra: medium/high | 30.1 | 46.1 | 13.3 | 33.3 |
|  | Ultra: median Ct value (IQR)<sup>&&</sup> | 20.3 (17.9 – 26.4) | 18.9 (17.5 – 23.3) | 25.0 (18.6 – 28.0) | 20.9 (17.9 – 28.3) |
|  | Culture-confirmed | 59.1 | 73.5 | 36.3 | 72.8 |
|  | Median days-to-culture-positivity (IQR) | 10.9 (7.1 – 16.9) | 9.8 (6.6 – 13.9) | 14.0 (6.8 – 17.8) | 11.8 (7.7 – 17.3) |
| Radiological status | Median CAD4TB v6 score (IQR) | 78 (49 – 97) | 89 (72 – 98) | 57 (47 – 87) | 77 (52 – 98) |
|  | Cavitation | 20.5 | 34.4 | 9.0 | 18.3 |
|  | Advanced extent of disease** | 39.0 | 57.3 | 23.1 | 38.0 |

PLHIV people living with HIV; ART antiretroviral treatment; IQR interquartile range; HDSS health and demographic surveillance system; WHO SS World Health Organization TB symptom screen; BMI body mass index; Ct cycle threshold; CAD4TB computer-aided detection of TB software
\* VL unknown but on ART for > 6 months and CD4 count > 200 (n=10)
& Microbiological status based on samples collected as part of study on day of treatment initiation other than programmatic (diagnostic) sample
&& Median of lowest Ct value for 4 *rpoB* probes given *M. tuberculosis* detected on at least one Ultra test
\*\* Defined as per National Tuberculosis Association (NTA) classification (NTA-III)<sup>40</sup>

The median age of child contacts was 9.1 years; 52% were male and 2% were living with HIV. Most were a child (58%) or sibling of the index person, and 65% had shared a room with them (Table S2).

### OUTCOME DATA

The overall prevalence of MTBC immunoreactivity among child contacts was 18% [138/755] (95% CI 15–22; cluster-adjusted using GEE). Intra-class correlation (ICC) was 0.38, indicating strong within-household clustering. MTBC immunoreactivity showed a strong dose-response relationship with the index person’s sputum MTBC bacillary burden (Figure 2).

**Figure 2.**
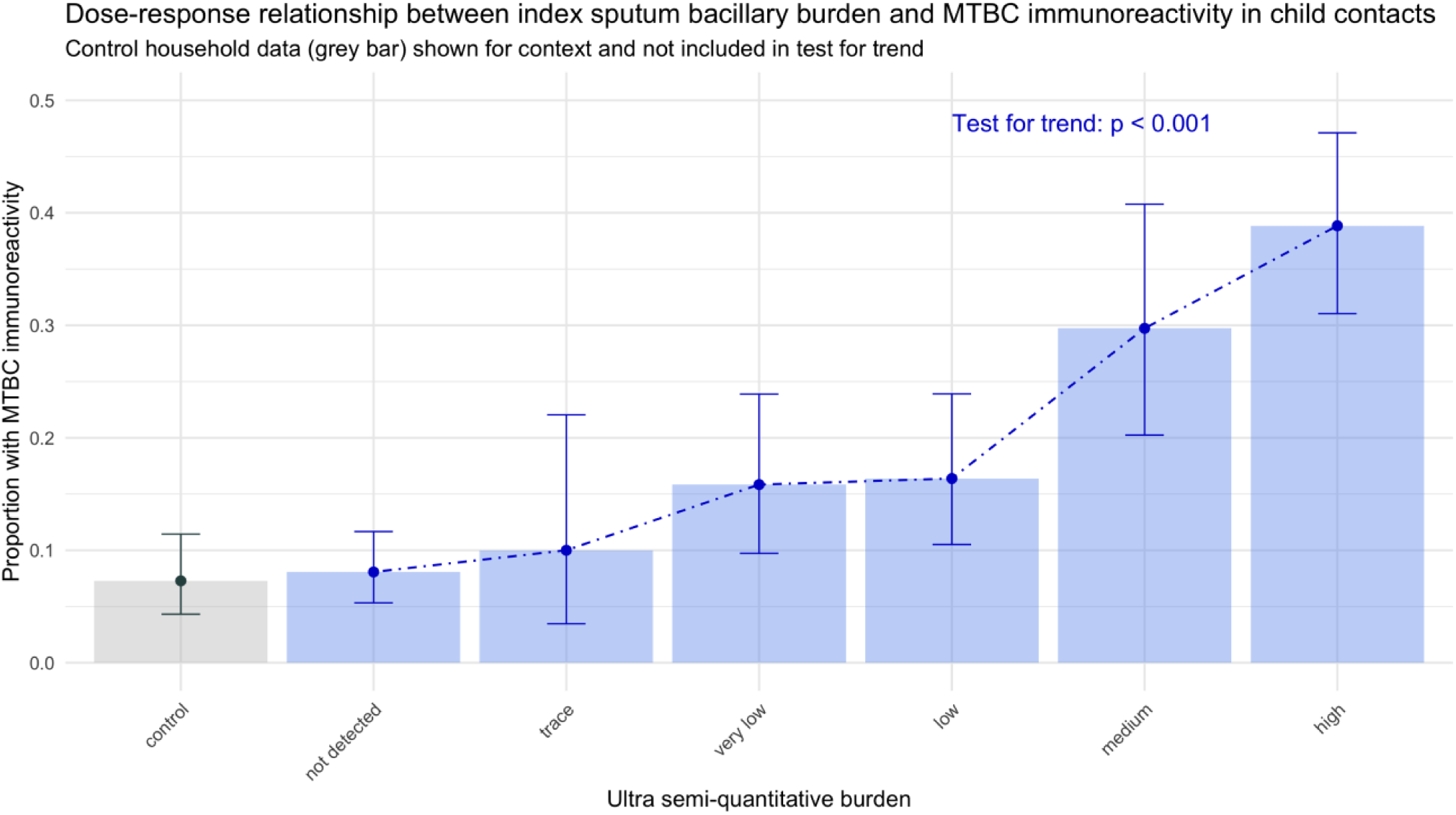
Dose-response relationship between index person sputum bacillary burden measured (Xpert MTB/RIF Ultra semi-quantitative output) and proportion of child contacts with MTBC immunoreactivity (bars represent cluster-adjusted 95% confidence intervals)

Index person-, child- and household-level factors associated with MTBC immunoreactivity are shown in Tables S3 and S4, respectively.

Among children in control households, immunoreactivity prevalence was 7% [15/206] (95% CI, 4–11; cluster-adjusted using GEE), with minimal within-household clustering (ICC=0.03).

### PRIMARY ANALYSIS

#### MTBC transmission risk from PLHIV not on effective ART versus those on effective ART

Eighty-one PLHIV not on effective ART and 113 PLHIV on effective ART were included in this analysis. Compared to PLHIV on effective ART, those not on effective ART were younger (median age 35 vs. 39 years), more frequently male (56.8% vs. 35.4%), and had more advanced immunosuppression (median CD4 count 174 vs. 561 cells/mm³) and more severe tuberculosis disease (Table 1). This was reflected in higher prevalence of WHO-defined symptoms (93% vs. 79%), lower BMI (<18.5; 19.8% vs. 10.6%), greater mycobacterial burden (33.3% vs. 13.3% with medium/high Ultra result), more frequent culture confirmation (72.8% vs. 36.3%), and a higher proportion with cavitary disease (18.3% vs. 9.0%).

Despite these differences, effective ART status was not associated with the risk of within-household MTBC transmission (TMLE risk ratio [RR] for no effective ART vs. effective ART, 1.07; 95% CI, 0.66 to 1.74; P=0.77; Table 2).

**Table 2.** Within-household MTBC transmission risk by ART status and symptom status estimated using TMLE.

| (1) ART status<br>(n = 467 contacts of 180 index PLHIV) | Number of<br>index<br>persons | Proportion child<br>contacts<br>QFT-positive (n/N) | Predicted probability of<br>MTBC immunoreactivity<sup>a</sup><br>(95% CI) | Risk ratio<sup>a</sup><br>(95% CI) | p-value |
| --- | --- | --- | --- | --- | --- |
| On effective ART | 103 | 0.147 (38/259) | 0.145 (0.094; 0.196) | 1 | 0.77 |
| Not on effective ART | 77 | 0.144 (30/208) | 0.156 (0.100; 0.212) | 1.07 (0.66 – 1.74) |  |
| (2) Symptom status<br>(n = 398 contacts of 157 index PLHIV engaged in<br>ART care) |  |  |  |  |  |
| At least one of the 4 WHO symptoms | 130 | 0.155 (52/335) | 0.151 (0.105; 0.198) | 1 | 0.47 |
| None of the 4 WHO symptoms | 27 | 0.095 (6/63) | 0.121 (0.055; 0.186) | 0.80 (0.43 – 1.47) |  |
| At least one symptom | 139 | 0.150 (53/354) | 0.146 (0.101; 0.191) | 1 | 0.27 |
| No symptoms | 18 | 0.113 (5/44) | 0.101 (0.042; 0.161) | 0.69 (0.36 – 1.33) |  |
| Cough of any duration | 96 | 0.184 (45/245) | 0.174 (0.121; 0.227) | 1 | 0.03 |
| No cough | 61 | 0.085 (13/153) | 0.106 (0.066; 0.145) | 0.61 (0.38 – 0.96) |  |

MTBC *Mycobacterium tuberculosis*; ART antiretroviral treatment; TMLE targeted minimum loss-based estimation; QFT QuantiFERON Gold plus assay; CI confidence interval
<sup>a</sup> Predicted probabilities and effect measures were estimated using TMLE with adjustment for *a priori* covariates (index age, index sex, geographical area, household socioeconomic status) based on directed acyclic graph

### SECONDARY ANALYSES

#### MTBC transmission risk of asymptomatic versus symptomatic PLHIV engaged in ART care

WHO symptom status among PLHIV engaged in ART care was not associated with MTBC transmission risk (adjusted RR 0.80, 95% CI 0.43–1.47). Similarly, the presence of any reported symptom versus none showed no evidence for any association (adjusted RR 0.69, 95% CI 0.36–1.33). However, index persons who did not report any cough had a 39% lower risk of transmission (adjusted RR 0.61, 95% CI 0.38–0.96; P=0.03) (Table 2).

#### MTBC transmission risk from HIV-negative versus PLHIV

Within-household transmission risk was greatest in HIV-negative women (30.5%; 95% CI, 25.9–35.2). Compared to HIV-negative women, women living with HIV had a 53% lower within-household MTBC transmission risk (14.3%; 95% CI, 11.2–17.4; RR 0.47; 95% CI 0.36-0.61). (Table 3). In contrast, within-household transmission risk did not differ between HIV-negative and HIV-positive men (12.5% and 12.6%, respectively; RR 1.01; 95% CI 0.71-1.42). Figure S6 illustrates the observed reciprocal effects of index person sex and HIV status on the risk difference and risk ratio scale.

**Table 3.**
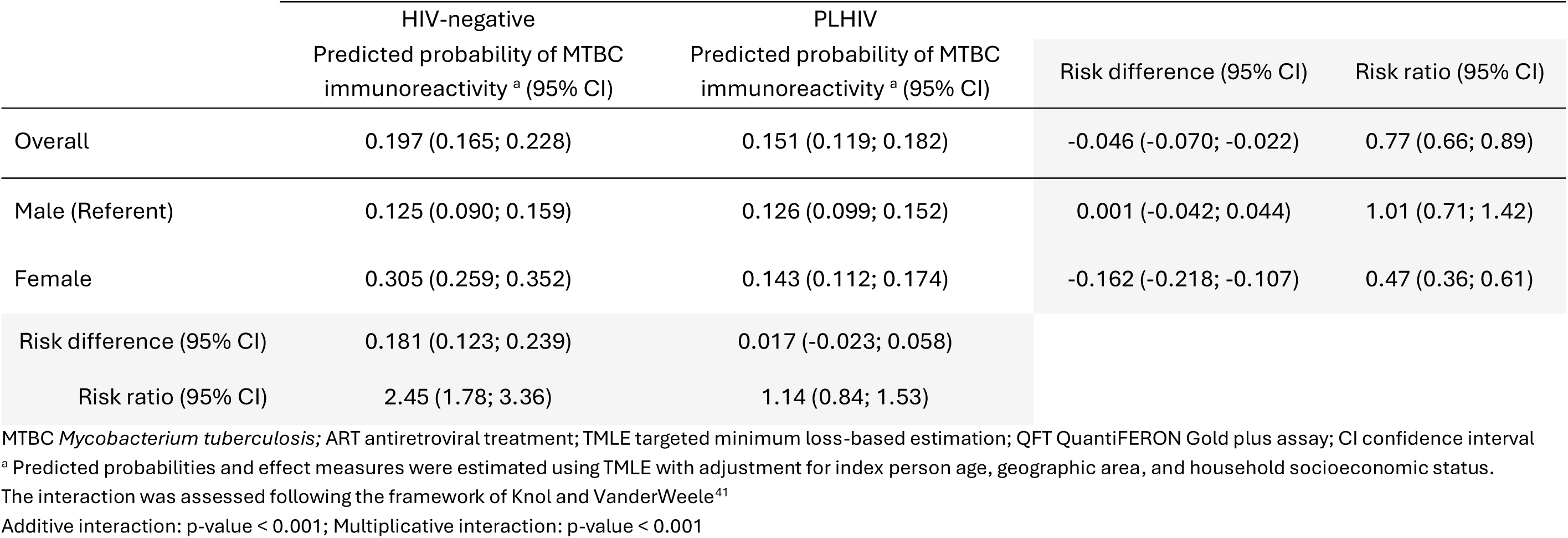
Interaction between HIV status and sex of index person on within-household MTBC transmission risk using TMLE.

|  | HIV-negative<br>Predicted probability of MTBC<br>immunoreactivity <sup>a</sup> (95% CI) | PLHIV<br>Predicted probability of MTBC<br>immunoreactivity <sup>a</sup> (95% CI) | Risk difference (95% CI) | Risk ratio (95% CI) |
| --- | --- | --- | --- | --- |
| Overall | 0.197 (0.165; 0.228) | 0.151 (0.119; 0.182) | -0.046 (-0.070; -0.022) | 0.77 (0.66; 0.89) |
| Male (Referent) | 0.125 (0.090; 0.159) | 0.126 (0.099; 0.152) | 0.001 (-0.042; 0.044) | 1.01 (0.71; 1.42) |
| Female | 0.305 (0.259; 0.352) | 0.143 (0.112; 0.174) | -0.162 (-0.218; -0.107) | 0.47 (0.36; 0.61) |
| Risk difference (95% CI) | 0.181 (0.123; 0.239) | 0.017 (-0.023; 0.058) |  |  |
| Risk ratio (95% CI) | 2.45 (1.78; 3.36) | 1.14 (0.84; 1.53) |  |  |

MTBC *Mycobacterium tuberculosis*; ART antiretroviral treatment; TMLE targeted minimum loss-based estimation; QFT QuantiFERON Gold plus assay; CI confidence interval
<sup>a</sup> Predicted probabilities and effect measures were estimated using TMLE with adjustment for index person age, geographic area, and household socioeconomic status.
The interaction was assessed following the framework of Knol and VanderWeele<sup>41</sup>
Additive interaction: p-value < 0.001; Multiplicative interaction: p-value < 0.001

#### Household-attributable risk of MTBC immunoreactivity: index versus control households

The household-attributable risk difference was 0.11 (95% CI 0.08 – 0.15), indicating an 11 percentage-point higher proportion of MTBC immunoreactivity among children living in TB-affected (index) households compared with children living in control households. This corresponded to a household-attributable fraction of 58.7% (95% CI 47.4 – 67.4%), indicating that nearly 60% of MTBC immunoreactivity among children living in TB-affected households was attributable to household exposure to the index person with pulmonary tuberculosis (Table S9).

## Discussion

We found no evidence for an association between within-household MTBC transmission risk and effective ART status or the WHO symptom status of index PLHIV. However, the absence of any cough was associated with substantially lower within-household MTBC transmission within households of index PLHIV engaged in ART care. Although there was no evidence for an association with alternative asymptomatic status definitions, their estimated effects were in the same direction, also suggesting reduced transmission risk.

Our most striking finding was the strong evidence for an interaction between within-household transmission risk and the index person’s HIV status and sex, whereby transmission risk to child contacts of HIV-negative women was more than twice that of women living with HIV, a disparity not seen among men. This observed biosocial interaction between index sex and HIV status suggests that neither biological nor social explanations will alone suffice to explain MTBC transmission.^24^ Women in many settings are primary caregivers and thus experience more prolonged and intimate contact with children, increasing the likelihood of MTBC exposure for the child. At the same time, HIV-negative women may have less engagement with health services than women with HIV, who often receive regular care, health education, and preventive interventions such as regular screening for tuberculosis and tuberculosis preventative treatment. This duality of greater exposure through social role and reduced mitigation through health system contact resulting in prolonged duration of infectiousness may explain the elevated within-household MTBC transmission risk we observed.

We found no association between effective ART status and within-household MTBC transmission risk (Table 2: RR, 1.07; 95% CI, 0.66 to 1.74; P=0.77). The theoretical schema in Figure 1 highlights the complexity of the impact of ART on MTBC transmission risk, which may operate through multiple, often opposing, pathways, with the net effect determined by which mechanisms predominate in a given population or setting. We postulate that PLHIV with excellent engagement in ART care likely undergo regular screening, leading to earlier tuberculosis diagnosis, reduced progression to severe, infectious disease, and a shortened period of infectiousness. This interpretation is supported by clinical differences at treatment initiation, where PLHIV not on effective ART appeared to have more advanced tuberculosis than those receiving ART. Direct measurement of the duration of infectiousness was not feasible, but its contribution could potentially be inferred through causal mediation analyses.

Historically, advanced HIV has been associated with limited MTBC transmission, driven by paucibacillary disease with minimal radiological involvement^25^ and high mortality that shortens the infectious period.^26^ We hypothesized that ART would narrow this gap by shifting the tuberculosis phenotype in PLHIV toward that observed in HIV-negative individuals. Yet, we still observed lower within-household MTBC transmission risk from PLHIV than from HIV-negative people (Table 3; RR, 0.77; 95% CI, 0.66 to 0.89), suggesting that meaningful differences in infectiousness persist despite effective HIV treatment. Taken together, these findings reinforce longstanding evidence that PLHIV are less infectious than their HIV-negative counterparts, although the underlying causal pathways may now differ.

Although reported symptom status may be an imperfect indicator of disease severity and infectiousness compared with objective measures such as cavitation and sputum bacillary burden,^27^ we observed a reduced risk of MTBC transmission in households where the index person did not report any cough (RR, 0.61; 95% CI, 0.38–0.96), supporting prior evidence of cough as a key driver of MTBC transmission.^28^

In a post hoc comparison of index and control households, the household-attributable risk difference was 0.11 (95% CI 0.08 to 0.15) and corresponding household-attributable fraction was 58.7% (95% CI 47.4–67.4; Table S10). That the majority of immunoreactivity among children in index households was attributable to household co-residence, rather than to the surrounding community, supports MTBC immunoreactivity as an informative marker of household transmission, despite the high community tuberculosis burden in this setting. This interpretation is strengthened by the strong dose–response relationship between the index person’s sputum bacillary burden and child immunoreactivity (Figure 2), which diffuse community exposure would not readily explain, and by the marked clustering of immunoreactivity within index households (ICC = 0.38) relative to control households (ICC = 0.03), consistent with a within-household source rather than community-acquired. Beyond informing future study design, the magnitude of this clustering may also offer insight into biological heterogeneity in transmissibility.

Our household-attributable fraction of 58.7% may appear to contrast with estimates that household exposure accounts for only 10–30% of *M. tuberculosis* transmission to children in high-burden settings.^29^ However, these quantities are not directly comparable. The latter is a population-attributable fraction, reflecting that most children in high-burden communities are not exposed to a known household index case and acquire infection (as measured in those studies) in the community; our estimate is an attributable fraction among children who do co-reside with a bacteriologically-confirmed case, quantifying the contribution of household co-residence conditional on exposure. The two are reconciled by the relative rarity of household exposure at the population level, and our findings are therefore consistent with the view that, although the household is an intense site of transmission for exposed children, community-acquired exposure predominates at the population scale. The substantial community-acquired background immunoreactivity evident in our control households reinforces this interpretation.

Tuberculosis household contact studies have long been used to quantify transmission risk, originating from Frost’s adaptation of Chapin’s concept of the secondary attack rate (SAR).^30^ The SAR captures key determinants of transmission, namely probability per contact, duration of infectiousness, exposure intensity, and contact susceptibility,^31^ but can be biased in high-incidence settings where transmission may also occur from community or undiagnosed household sources.^32,33^ By measuring MTBC immunoreactivity in children from control households without tuberculosis, we were able to contextualize observed transmission patterns and account, in part, for background community exposure.

Contact intensity is a key determinant of within-household MTBC transmission,^32,34^ but lies on the causal pathway between our exposures and transmission risk; accordingly, it was not adjusted for. To disentangle these pathways, we plan causal mediation analyses, extending methods from prior studies.^9,35^ These approaches allow evaluation of other intermediaries, such as bacillary burden, cavitation, and novel radiologic markers like CAD4TB, and enable decomposition of mediation and interaction effects.^36^ Though methodologically complex, they are essential to elucidate how clinical, biological, and social factors intersect to drive transmission.

Our findings also contribute to the broader debate on interaction.^24,37,38^ Pure interaction (a special form of qualitative interaction)^37^ suggests that the observed effect modification is not merely statistical, but reflects an interdependence^36^ between HIV status and sex of the index person in shaping within-household MTBC transmission risk. This likely reflects both biological and social mechanisms, including differences in bacillary burden, disease extent, infectious period, and gendered patterns of health-seeking and contact, warranting further study in settings with lower HIV prevalence.

Several limitations warrant consideration. The sample size was smaller than intended because of unavoidable delays during the COVID-19 pandemic. More fundamentally, a positive QFT result indicates immune sensitisation to MTBC rather than necessarily active or recent infection and does not identify the timing or source of exposure; in a setting with ongoing community transmission, a proportion of immunoreactivity will inevitably reflect community-acquired rather than household exposure. MTBC immunoreactivity is therefore best understood as a marker of cumulative MTBC exposure resulting in immune sensitisation, with our comparison against geographically matched control households serving to isolate the component of that exposure attributable to index-household co-residence. Viewed this way, a household-attributable fraction approaching 60%, sustained above an active community background, likely approaches the upper limit of what a cross-sectional marker of immunoreactivity can attribute to the household. To minimise community-related misclassification, we restricted analyses to child contacts, who are less likely to have had extensive community exposure, and incorporated HIV testing of children to reduce bias from false-negative immunoreactivity, although HIV prevalence in this group was very low. Alternative designs, such as longitudinal QFT testing or pathogen-level linkage between index and secondary infections, could refine attribution of source and timing, but each carries its own constraints, including residual uncertainty about the source of incident infection and reliance on progression to culturable disease, which occurs in only a minority of those infected. Finally, because index persons were enrolled through primary healthcare facilities, our assessment of asymptomatic tuberculosis was limited to PLHIV attending ART care and may not capture the broader spectrum of asymptomatic tuberculosis in the community.

Despite the issues, household studies remain a powerful tool for understanding the mechanisms of MTBC transmission.^39^ Households are the highest-risk environment due to prolonged, close-proximity contact, even if they do not reflect the more casual and sporadic interactions typical of community-level transmission. Accordingly, while household studies cannot fully describe population-level transmission, they provide essential insight into the drivers of spread and the characteristics of individuals most likely to transmit infection.

## Conclusions

In this observational study, we found no evidence that effective ART or WHO4SS status influenced within-household MTBC transmission risk, although wide confidence intervals limit firm conclusions. The absence of cough in the index person was, however, associated with reduced risk and within-household transmission risk was highest among child contacts of HIV-negative women. These findings suggest that reported cough is a useful marker of transmission risk and that routine tuberculosis screening within ART care may reduce transmission from PLHIV; intensified efforts are nonetheless needed to achieve earlier tuberculosis detection in HIV-negative individuals.

## Data Availability

The data underlying this article will be made available online at LSHTM Data Compass (https://datacompass.lshtm.ac.uk/) upon publication

## NOTES

This research was supported by funding from the National Institute of Allergy and Infectious Diseases of the National Institutes of Health under award number: 1R01AI147321-01. The content is solely the responsibility of the authors and does not necessarily represent the official views of the National Institutes of Health.

We thank the men, women and children who participated in this study; the Department of Health staff at the healthcare facilities; Evashni Rampersad for administrative support; Kathy Baisley (PhD) for statistical advice; the AHRI field and data management teams and AHRI Clinical Laboratory staff who supported this study.

For the purposes of open access, the authors have applied a Creative Commons Attribution (CC BY) licence to any Accepted Author Manuscript version arising from this submission.

## Notes

### Competing Interest Statement

The authors have declared no competing interest.

### Clinical Protocols

https://wellcomeopenresearch.org/articles/9-622

### Author Declarations

The study was approved by ethics committees at the University of KwaZulu-Natal (BF472/19) and the London School of Hygiene & Tropical Medicine, (17728-6).

## References

1. WHO. Global tuberculosis control: WHO report 2024 [Internet]. 2024 [cited 2025 Apr 21];Available from: https://www.who.int/publications/i/item/9789240101531

2. WHO. Global tuberculosis control: WHO report 2022 [Internet]. 2022 [cited 2025 Apr 21];Available from: https://www.who.int/publications/i/item/9789240061729

3. Dye C, Williams BG. Tuberculosis decline in populations affected by HIV: a retrospective study of 12 countries in the WHO African Region. Bull World Health Organ 2019;97(6):405–14.

4. Elliott AM, Hayes RJ, Halwiindi B, et al. The impact of HIV on infectiousness of pulmonary tuberculosis: a community study in Zambia. AIDS 1993;7(7):981–7.

5. Cauthen GM, Dooley SW, Onorato IM, et al. Transmission of Mycobacterium tuberculosis from tuberculosis patients with HIV infection or AIDS. Am J Epidemiol 1996;144(1):69–77.

6. Espinal MA, Perez EN, Baez J, et al. Infectiousness of Mycobacterium tuberculosis in HIV-1-infected patients with tuberculosis: a prospective study. Lancet 2000;355(9200):275–80.

7. Carvalho AC, DeRiemer K, Nunes ZB, et al. Transmission of Mycobacterium tuberculosis to contacts of HIV-infected tuberculosis patients. Am J Respir Crit Care Med 2001;164(12):2166–71.

8. Kenyon TA, Creek T, Laserson K, et al. Risk factors for transmission of Mycobacterium tuberculosis from HIV-infected tuberculosis patients, Botswana. Int J Tuberc Lung Dis 2002;6(10):843–50.

9. Huang CC, Tchetgen ET, Becerra MC, et al. The effect of HIV-related immunosuppression on the risk of tuberculosis transmission to household contacts. Clin Infect Dis Off Publ Infect Dis Soc Am 2014;58(6):765–74.

10. Martinez L, Shen Y, Mupere E, Kizza A, Hill PC, Whalen CC. Transmission of Mycobacterium Tuberculosis in Households and the Community: A Systematic Review and Meta-Analysis. Am J Epidemiol 2017;185(12):1327–39.

11. Borgdorff MW, De Cock KM. Provision of ART to individuals infected with HIV: impact on the epidemiology and control of tuberculosis. Int J Tuberc Lung Dis 2017;21(11):1091–2.

12. Gareta D, Baisley K, Mngomezulu T, et al. Cohort Profile Update: Africa Centre Demographic Information System (ACDIS) and population-based HIV survey. Int J Epidemiol 2021;50(1):33–4.

13. Wong EB, Olivier S, Gunda R, et al. Convergence of infectious and non-communicable disease epidemics in rural South Africa: a cross-sectional, population-based multimorbidity study. Lancet Glob Health 2021;9(7):e967–76.

14. Africa Health Research Institute (AHRI) Household Contact study: a … [Internet]. Wellcome Open Res. Open Access Publ. Platf. 2024 [cited 2025 Apr 21];Available from: https://wellcomeopenresearch.org/articles/9-622#ref-1

15. Getahun H, Kittikraisak W, Heilig CM, et al. Development of a standardized screening rule for tuberculosis in people living with HIV in resource-constrained settings: individual participant data meta-analysis of observational studies. Plos Med 2011;8(1):e1000391.

16. Lendle SD, Schwab J, Petersen ML, Laan MJ van der. ltmle: An R Package Implementing Targeted Minimum Loss-Based Estimation for Longitudinal Data. J Stat Softw 2017;81:1–21.

17. Rose S, van der Laan MJ. LTMLE [Internet]. In: van der Laan MJ, Rose S, editors. Targeted Learning in Data Science: Causal Inference for Complex Longitudinal Studies. Cham: Springer International Publishing; 2018 [cited 2023 Sept 22]. p. 35–47.Available from: 10.1007/978-3-319-65304-4_4

18. GitHub - benoitlepage/MargIntTmle: Marginal interaction effects by g-computation, IPTW or TMLE [Internet]. [cited 2025 May 5];Available from: https://github.com/benoitlepage/MargIntTmle/tree/main

19. VanderWeele TJ. Sufficient Cause Interactions and Statistical Interactions. Epidemiology 2009;20(1):6.

20. Lash TL, VanderWeele TJ, Haneause S, Rothman KJ. Modern epidemiology. Fourth edition. Philadelphia: Lippincott Williams & Wilkins; 2021.

21. CRAN: geepack citation info [Internet]. [cited 2025 Apr 13];Available from: https://cran.r-project.org/web/packages/geepack/citation.html

22. Arel-Bundock V, Greifer N, Heiss A. How to Interpret Statistical Models Using marginaleffects for R and Python. J Stat Softw 2024;111:1–32.

23. Smith MJ, Mansournia MA, Maringe C, et al. Introduction to computational causal inference using reproducible Stata, R, and Python code: A tutorial. Stat Med 2022;41(2):407–32.

24. Harris KM, McDade TW. The Biosocial Approach to Human Development, Behavior, and Health Across the Life Course. Russell Sage Found J Soc Sci RSF 2018;4(4):2–26.

25. Munthali L, Khan PY, Mwaungulu NJ, et al. The effect of HIV and antiretroviral therapy on characteristics of pulmonary tuberculosis in northern Malawi: a cross-sectional study. BMC Infect Dis 2014;14:107.

26. Corbett EL, Bandason T, Cheung YB, et al. Prevalent infectious tuberculosis in Harare, Zimbabwe: burden, risk factors and implications for control. Int J Tuberc Lung Dis 2009;13(10):1231–7.

27. McCreesh N, MacPherson P, Bampi J, Engel N, Kranzer K, Khan P. Reported tuberculosis symptoms: an inadequate classifier of disease state [Internet]. 2025 [cited 2025 Aug 28];Available from: https://osf.io/fhpeu_v1

28. Turner RD, Bothamley GH. Cough and the Transmission of Tuberculosis. J Infect Dis 2014;211(9):1367–72.

29. Martinez L, Lo NC, Cords O, et al. Paediatric tuberculosis transmission outside the household: challenging historical paradigms to inform future public health strategies. Lancet Respir Med 2019;7(6):544–52.

30. Frost WH. Risk of Persons in Familial Contact with Pulmonary Tuberculosis. Am J Public Health Nations Health 1933;23(5):426–32.

31. Yates TA, Khan PY, Knight GM, et al. The transmission of Mycobacterium tuberculosis in high burden settings. Lancet Infect Dis 2016;16(2):227–38.

32. Morozova O, Cohen T, Crawford FW. Risk ratios for contagious outcomes. J R Soc Interface 2018;15(138):20170696.

33. Kendall EA. When Infections Don’t Reflect Infectiousness: Interpreting Contact Investigation Data With Care. Clin Infect Dis 2020;73(9):e3456–8.

34. Layan M, Hens N, de Hoog MLA, Bruijning-Verhagen PCJL, Cowling BJ, Cauchemez S. Addressing current limitations of household transmission studies by collecting contact data. Am J Epidemiol 2024;193(12):1832–9.

35. Martinez L, Sekandi JN, Castellanos ME, Zalwango S, Whalen CC. Infectiousness of HIV Seropositive Tuberculosis Patients in a High-burden African Setting. Am J Respir Crit Care Med 2016;194(9).

36. VanderWeele TJ. A Unification of Mediation and Interaction: A 4-Way Decomposition. Epidemiology 2014;25(5):749.

37. VanderWeele TJ, Knol MJ. A Tutorial on Interaction. Epidemiol Methods [Internet] 2014 [cited 2025 Jan 6];3(1). Available from: https://www.degruyter.com/document/doi/10.1515/em-2013-0005/html

38. Bours MJL. Tutorial: A nontechnical explanation of the counterfactual definition of effect modification and interaction. J Clin Epidemiol 2021;134:113–24.

39. Hill PC, Ota MO. Tuberculosis case-contact research in endemic tropical settings: design, conduct, and relevance to other infectious diseases. Lancet Infect Dis 2010;10(10):723–32.

40. NTA. Diagnostic Standards and Classification of Tuberculosis [Internet]. National Tuberculosis Association; 1940. Available from: https://catalog.hathitrust.org/Record/001562842

41. Knol MJ, VanderWeele TJ. Recommendations for presenting analyses of effect modification and interaction. Int J Epidemiol 2012;41(2):514–20.

