## Supplementary material for "Within-household transmission risk of pulmonary tuberculosis in the era of universal antiretroviral therapy"

Khan PY et al

### Supplementary Methods

#### Laboratory procedures for QFT testing

To minimize measurement error due to technical variability of the QuantiFERON-TB Gold (QFT) assay (Qiagen, Hilden, Germany),[1,2] we adhered to a strict protocol encompassing standardized handling of whole-blood specimens, time to incubation, incubation times, and interpretation criteria for defining a positive assay, which was used to determine MTBC immunoreactivity.

During the home visits, between 4 and 6 mL of blood was collected from participating children into a single lithium heparin tube, mixed according to the manufacturer's instructions, and transported to the laboratory at room temperature. Within 4 hours of collection, 1 mL of blood was inoculated into each of the four QFT-Plus tubes and incubated at 37 ± 1°C for 20 hours. After incubation, tubes were centrifuged for 15 minutes at 2000–3000 × g, and plasma was harvested and stored at –80°C until testing. The standard QFT-Plus enzyme-linked immunosorbent assay (ELISA) kit was used. The optical density of each well was measured with a plate reader (BioTek 800TS), and plate validation and result interpretation were configured using the microplate reader software (BioTek Gen5). The concentration of released interferon-gamma (IFN-γ) in each tube was calculated by subtracting the value of the nil (negative control) tube. Assays with a coefficient of variation below 15% and a standard curve correlation coefficient above 0.98 were deemed technically valid. All results were interpreted with reference to a 4-point standard curve, as specified by the manufacturer.

#### Causal identifiability assumptions for primary analysis

For the primary aim, our target causal parameter was defined by the contrasts in the counterfactual prevalence of MTBC immunoreactivity in child contacts of index PLHIV not on effective ART at the time of treatment initiation compared to child contacts of index PLHIV on effective ART at the time of treatment initiation.

To support a valid causal interpretation using an observational study design, we needed assumptions external to the data or so-called identifying assumptions.[3] In general, conditions for nonparametric identification of average causal effects include no interference, consistency, conditional exchangeability and positivity.[4] For the purposes of this observational study, we assumed:

(1) *No interference*

We assumed that ART status of the index PLHIV with TB was unlikely to affect the QFT-status of a child living in another index person's household. Only 12/227 (5.3%) index persons with TB (PWTB) lived within 100 metres of another index PWTB in our study.

(2) *Consistency*

Our hypothetical intervention (exposure) was well-defined in that PLHIV had been on ART for at least 6 months and had to have a fully suppressed HIV viral load (<50 copies/ml). However, the stage of HIV infection at the time of starting ART treatment may be an issue for consistency of the effect of "effective ART" on the risk of intra-household transmission in that PLHIV who started ART earlier during their HIV infection are different immunologically (and behaviorally) to PLHIV who were diagnosed at a more much advanced stage of HIV.

(3) *Conditional exchangeability*

This was an unverifiable assumption of no unmeasured confounding after we have adjusted for index age, index sex, household SES and residential area (our *a priori* confounders).

(4) *Positivity*

There was good overlap in propensity scores (Probability(exposure | confounders)) confirming that there are no positivity violations (see Figure S5 in Supplementary Figures).

#### Variable definitions

Outcome

The outcome was MTBC immunoreactivity, defined as positive QFT result (immune sensitisation following exposure to MTBC antigens) in child household contacts (aged 2 – 14 years), used as a proxy for within-household MTBC transmission.

MTBC immunoreactivity provides a (proxy) measure of within-household transmission risk on the strong assumption that the index PWTB is the source of MTBC immunoreactivity in the child household contact. Restricting household contacts to children under 14 years minimises misclassification of within-household transmission risk resulting from an extra-household source as older adolescents and young adults are likely to have much higher risk of exposure to MTBC from contact outside of the household compared to younger children with limited social contacts. Excluding the youngest children aged under 2 years reduces the risk of an indeterminate assay result (missingness of outcome).

Exposures of interest

(1) On effective ART at TB treatment initiation defined as on ART for at least 6 months with a HIV VL < 50 copies/ml (binary variable). This analysis was restricted to PLHIV
(2) No WHO TB symptoms (cough, weight loss, fever and night sweats) reported at TB treatment initiation. This analysis was restricted to PLHIV engaged in ART care or starting ART as this group of clinic attendees are screened using sputum TB nucleic acid amplification (NAAT) testing irrespective of whether they report symptoms or not
(3) Joint (“synergistic”) effect of HIV status and sex of index PWTB. All index PWTB were included in this analysis

*Ascertainment of symptom status*: all index PWTB were asked the WHO-recommended four symptom screen (WHO4SS) of cough (of any duration), weight loss, night sweats and fever, followed by an open-ended question of whether they had any other symptoms to report. If the answer to this question was no, then no further questions were asked. If the answer is yes, participants were asked to specify which other symptoms they have from the following list: haemoptysis, chest pain, difficulty in breathing, loss of appetite, tiredness, nausea, vomiting, diarrhoea, abdominal pain, headache, back pain, joint pain, or lymphadenopathy, with capture of ‘other’ symptoms as free text.

Key confounders

*Index person level*

Age (continuous variable) of index PWTB

Sex (binary variable) of the index PWTB

*Household-level*

Socioeconomic status (ordinal variable: 4 categories)

A polychoric dual-component wealth index provides a measure of household socioeconomic status and was created using (i) ordered categorical (ordinal) variables, (ii) squared multiple correlations to remove variables weakly correlated with asset-based wealth to reduce noise in the principal component analysis (PCA), which uses (iii) polychoric correlations instead of Pearson's correlation and (iv) two principal components instead of one. This approach reduces the "urban bias" of the standard approach to generating a wealth index using information on assets, household characteristics and access to amenities resulting in a better representation of typically rural characteristics of wealth.[5]

Area of residence within the Africa Health Research Institute Health and Demographic Surveillance System (nominal variable: 3 categories – North, West and South).

Area of residence provides a proxy variable to account for community MTBC transmission. Index PWTB recruited from Mtubatuba, KwaMsane, Mpukunyoni, Madwaleni and Siphu Zungu healthcare facilities are categorised as resident in the "West"; index PWTB recruited from Nkundusi, Ntondweni, Hluhluwe and Hlabisa Hospital/Hlabisa Gateway healthcare facilities are categorised as resident in the "North" and index PWTB recruited from Somkhele, Esiyembeni, Machibini and Gunjaneni are categorised as resident in the "West". See Figure S1 for map of the study area showing location of healthcare facilities and underlying population distribution of the area. The first author undertook an analysis using 2023 TIER.net data (electronic TB treatment register[6] to provide number of microbiologically-confirmed PTB registrations aggregated by facility) and population count data from WorldPop[7] hub to estimate TB notifications per 100,000 population as a crude proxy measure of TB incidence by area. Results shown in Table S9 – these estimates were used to stratify AHRI HDSS into high transmission and low transmission areas used as part of a robustness check of the post-hoc analysis described below.

#### Household versus community-acquired risk of child MTBC immunoreactivity (post-hoc analysis)

This analysis was restricted to index person households within the AHRI HDSS with a matched control household. Using the same TMLE framework as the primary analysis (*ltmle* package in R), with the same ensemble machine-learning library and accounting for household-level clustering, we adjusted for child age and sex, household crowding (number of persons per sleeping room), and geographical area. Area was retained in the adjustment set despite the 1 km matching to address any residual spatial confounding. We estimated the causal risk difference, which is the difference in expected QFT-positivity had all children resided in index versus control households, and the corresponding attributable fraction, the proportion of expected QFT-positivity were every child to reside in an index household attributable to index-household co-residence, relative to the counterfactual in which every child instead resided in a geographically matched control household

### Supplementary Figures

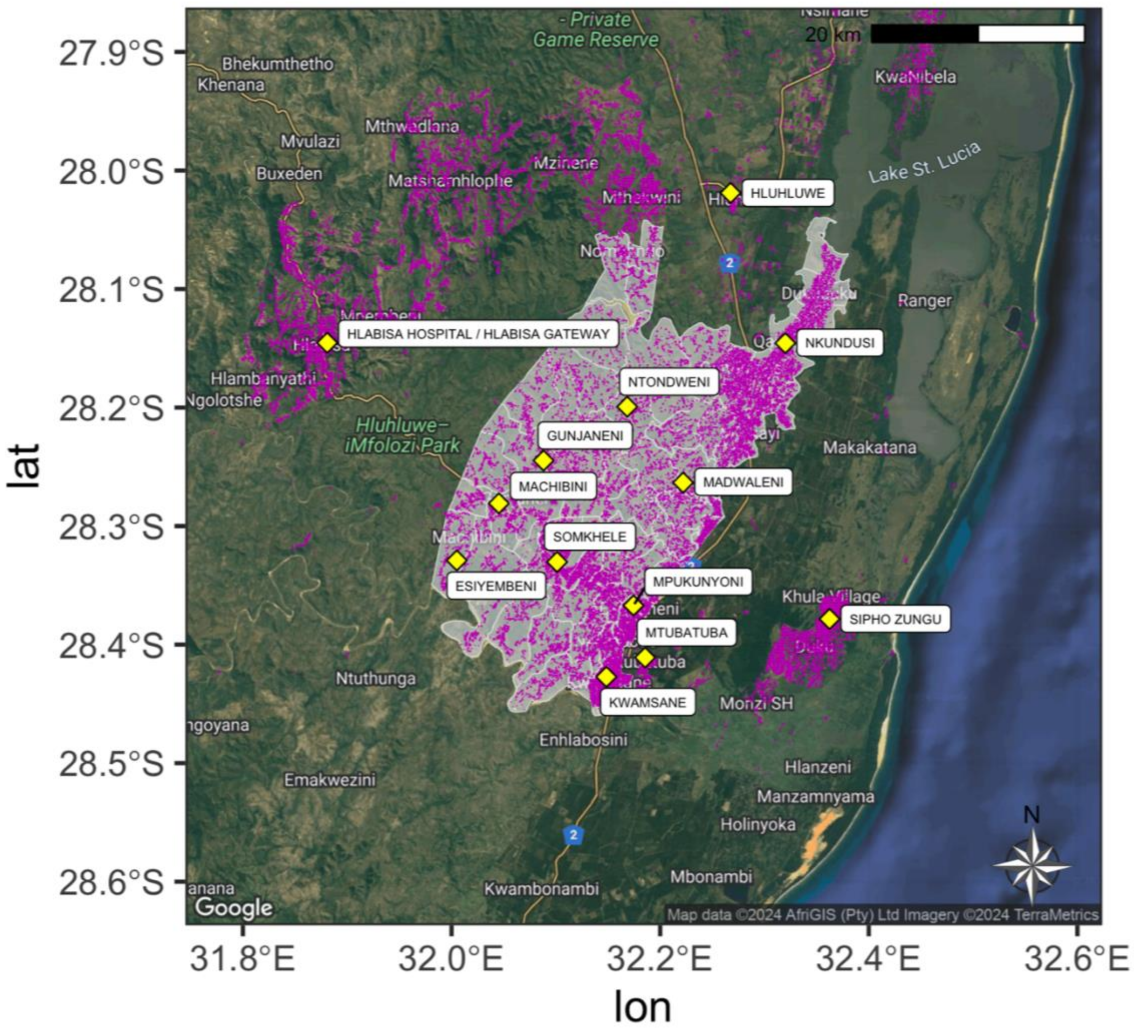

#### Figure S1. Map of study area showing population distribution within AHRI HDSS (purple shaded area).

The population data for 2020 for South Africa displayed in the map was obtained from WorldPop Hub (*https://hub.worldpop.org/geodata/listing?id=69*) which is also shared under the Creative Commons Attribution 4.0 International License.[7]

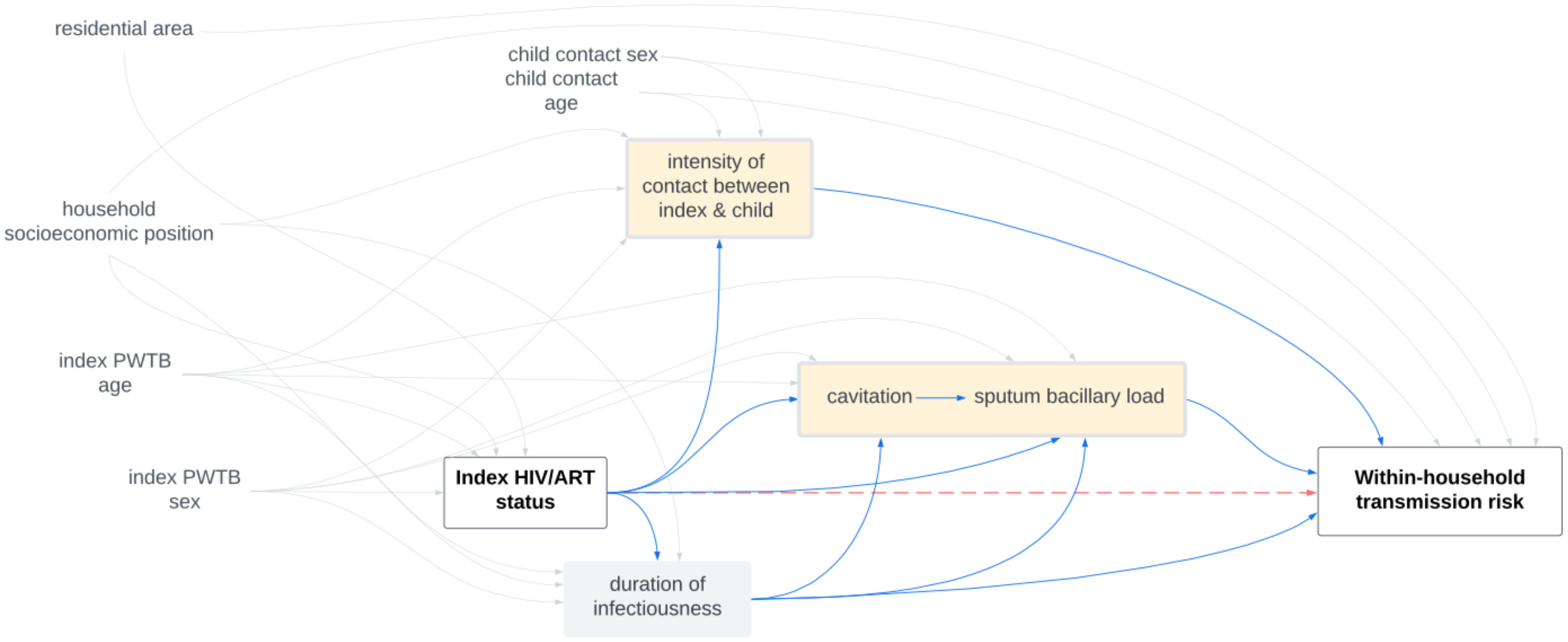

#### Figure S2. Causal diagram illustrating the structural assumptions of the underlying data-generating process of the relationship between HIV/ART status of index person with tuberculosis and risk of within-household MTBC transmission

Red dashed arrow highlights focal relationship between exposure of interest and outcome. Blue arrows highlight covariates (mediators – seen in peach boxes) on the causal pathway between exposure of interest and the outcome. Duration of infectiousness is the only unobserved covariate (grey box) in the causal diagram.

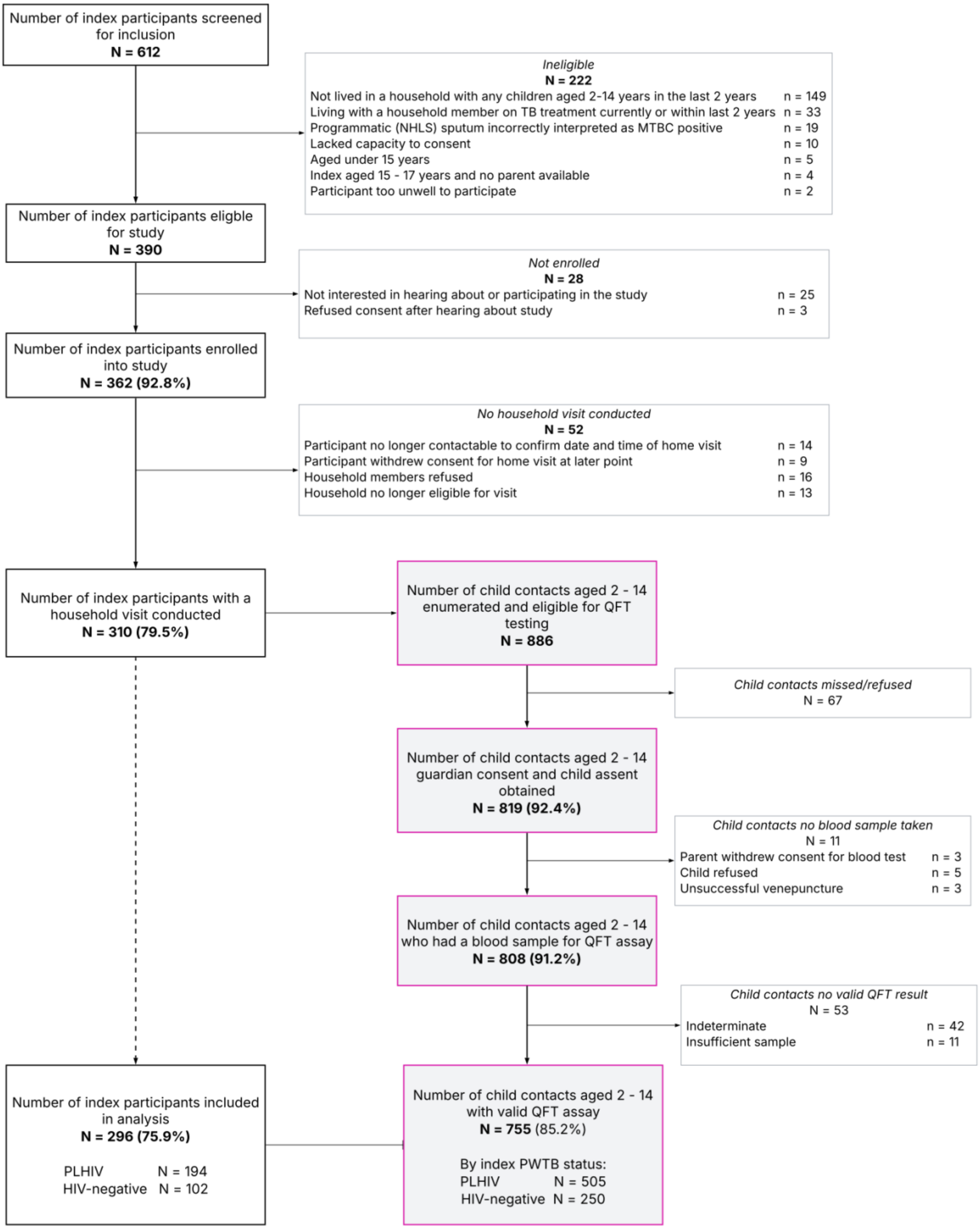
Number of index participants screened for inclusion
N = 612
Ineligible
N = 222
Not lived in a household with any children aged 2-14 years in the last 2 years n = 149
Living with a household member on TB treatment currently or within last 2 years n = 33
Programmatic (NHLS) sputum incorrectly interpreted as MTBC positive n = 19
Lacked capacity to consent n = 10
Aged under 15 years n = 5
Index aged 15 - 17 years and no parent available n = 4
Participant too unwell to participate n = 2
Number of index participants eligible for study
N = 390
Not enrolled
N = 28
Not interested in hearing about or participating in the study n = 25
Refused consent after hearing about study n = 3
Number of index participants enrolled into study
N = 362 (92.8%)
No household visit conducted
N = 52
Participant no longer contactable to confirm date and time of home visit n = 14
Participant withdrew consent for home visit at later point n = 9
Household members refused n = 16
Household no longer eligible for visit n = 13
Number of index participants with a household visit conducted
N = 310 (79.5%)
Number of child contacts aged 2 - 14 enumerated and eligible for QFT testing
N = 886
Child contacts missed/refused
N = 67
Number of child contacts aged 2 - 14 guardian consent and child assent obtained
N = 819 (92.4%)
Child contacts no blood sample taken
N = 11
Parent withdrew consent for blood test n = 3
Child refused n = 5
Unsuccessful venepuncture n = 3
Number of child contacts aged 2 - 14 who had a blood sample for QFT assay
N = 808 (91.2%)
Child contacts no valid QFT result
N = 53
Indeterminate n = 42
Insufficient sample n = 11
Number of index participants included in analysis
N = 296 (75.9%)
PLHIV N = 194
HIV-negative N = 102
Number of child contacts aged 2 - 14 with valid QFT assay
N = 755 (85.2%)
By index PWTB status:
PLHIV N = 505
HIV-negative N = 250

#### Figure S3. Study flowchart of index participants from eligibility to child household contacts tested for evidence of MTBC immunoreactivity

MTBC = *Mycobacterium tuberculosis complex*; NHLS = South Africa National Health Laboratory Service; PLHIV = people living with HIV; PWTB = people with TB; QFT = QuantiFERON Gold plus

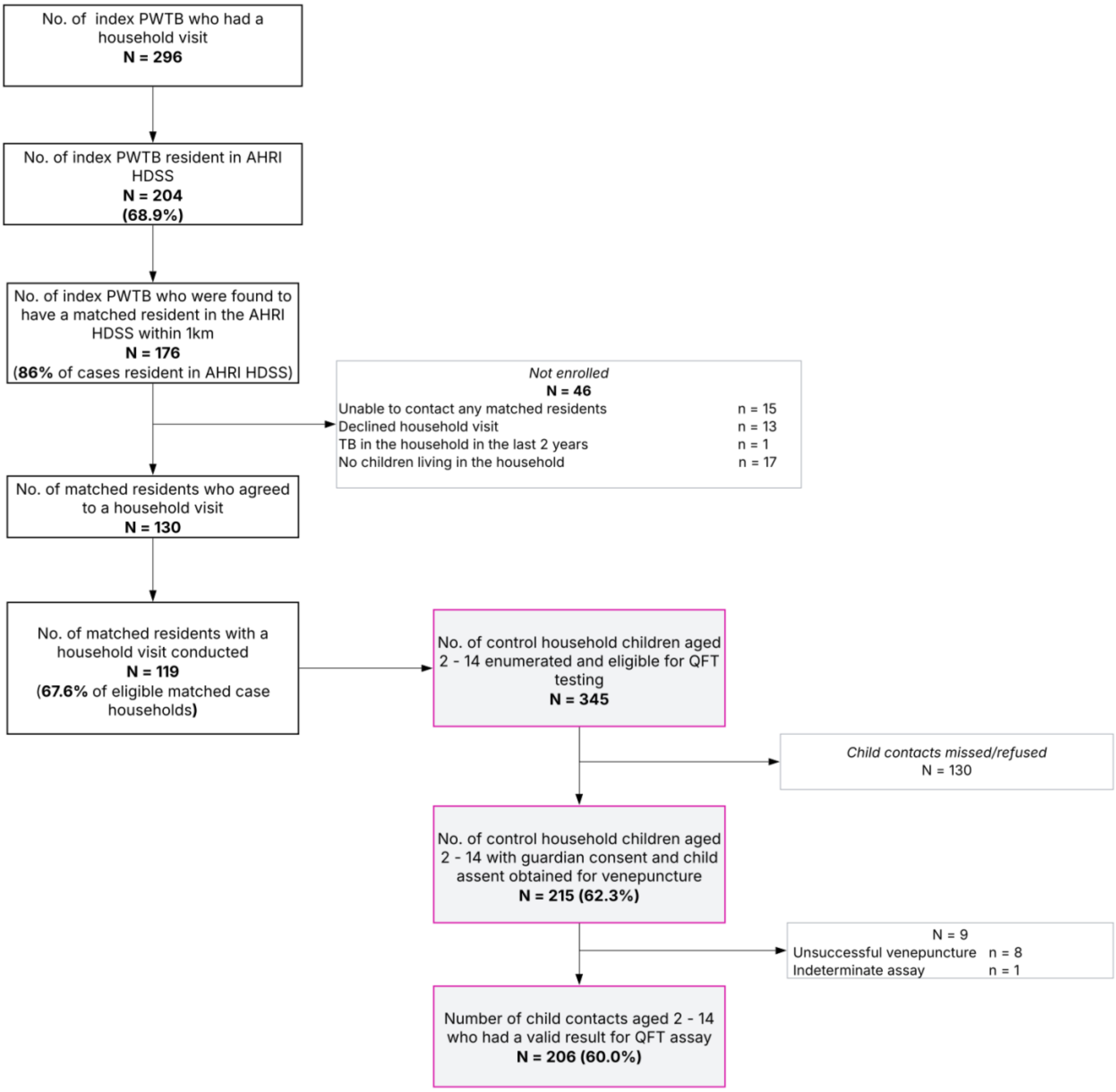

#### Figure S4. Study flowchart of matched control households from eligibility to testing of children tested for evidence of MTBC immunoreactivity

MTBC = *Mycobacterium tuberculosis complex*; NHLS = South Africa National Health Laboratory Service; PLHIV = people living with HIV; PWTB = people with TB; QFT = QuantiFERON Gold plus

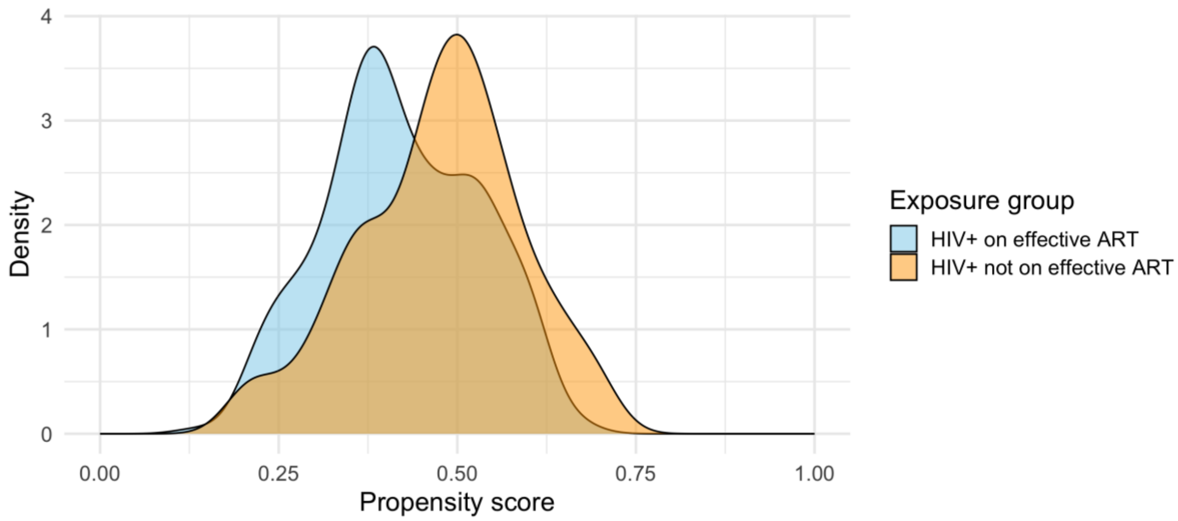

#### Figure S5. Propensity score distributions by ART status

To evaluate the assumption of positivity (see Supplementary Methods), we examined the distribution of estimated propensity scores across exposure groups. Figure S5 shows kernel density plots of propensity scores for participants who were HIV-positive on effective ART and those who were HIV-positive but not on effective ART. The distributions demonstrate good overlap between groups, with no evidence of sparsity at the extremes. This indicates that, across the range of covariate profiles, participants in both exposure groups had non-zero probabilities of assignment, supporting the validity of the positivity assumption for causal inference.

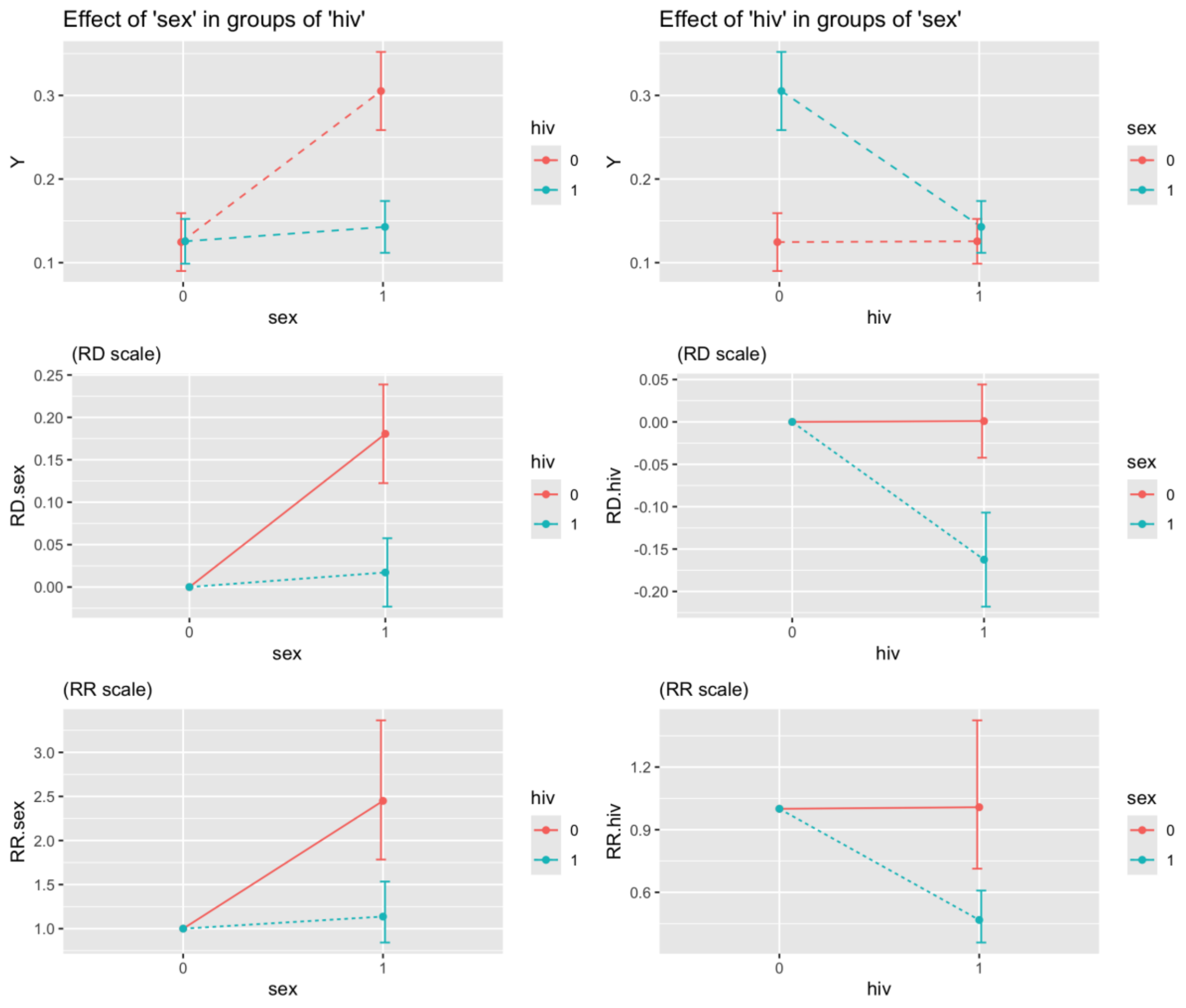

Figure S6. Interaction effects of index person HIV status and sex on within-household transmission risk estimated by TMLE (shown as predicted probabilities, on risk difference scale and on risk ratio scale)

RD = risk difference; RR = risk ratio; TMLE = targeted minimum loss-based estimation; Y = predicted probability of MTBC immunoreactivity

This figures clearly demonstrates the interaction between index person HIV status and sex on the risk difference and risk ratio scale. The first column shows that there is a higher predicted probability of MTBC immunoreactivity in child contacts of HIV-negative women compared to HIV-negative men whilst there is little difference between men and women living with HIV(effect of sex within HIV groups). The second column shows that there is a much lower predicted probability of MTBC immunoreactivity in contacts of women living with HIV compared to HIV-negative women whilst there is little difference between HIV-negative men and men living with HIV (effect of HIV within sex groups).

### Supplementary Tables

#### Table S1. Characteristics of index participants at enrolment by child contact data availability

| | | All index participants enrolled N = 362 col % | Index participants included in analyses N = 296 col % | Index participants without child contact data N = 66 col % |
|---|---|---|---|---|
| Age (years) | Median (IQR) | 36 (29 – 48) | 36 (29 – 47) | 37 (27 – 53) |
| Gender | Male (%) | 178 (49) | 147 (50) | 31 (47) |
| Education | Grade 12 and above | 153 (42) | 126 (43) | 27 (41) |
| Employment status | Unemployed | 282 (78) | 230 (78) | 52 (79) |
| Smoking status | Smoked in last 12 months | 72 (20) | 60 (20) | 12 (18) |
| Geographical area in HDSS | East | 161 (44) | 126 (43) | 35 (53) |
| | North | 97 (27) | 87 (29) | 10 (15) |
| | West | 104 (29) | 83 (28) | 21 (32) |
| HIV status | HIV- | 129 (36) | 102 (34) | 27 (41) |
| | HIV+ | 233 (64) | 194 (66) | 39 (59) |
| | ART started in last 6 months | 54/233 (23) | 45/194 (23) | 9/39 (23) |
| PLHIV | HIV viral load <50 copies/ml | 133/233 (57) | 110/194 (57) | 23/39 (59) |
| | Median CD4 count (IQR) | 452 (183 – 667) | 456 (190 – 640) | 423 (180 – 879) |
| Definition of asymptomatic TB | No symptoms | 30 (8) | 21 (7) | 9 (14) |
| | Negative on WHO SS | 50 (14) | 35 (12) | 15 (23) |
| | No cough | 112 (31) | 90 (30) | 22 (33) |
| Previous history of TB | | 87 (24) | 76 (26) | 11 (17) |
| BMI median (IQR) | | 22.0 (19.5 – 26.2) | 22.0 (19.5 – 26.4) | 22.2 (19.5 – 25.1) |
| Microbiological status[§] | Diagnostic (programmatic) Ultra trace | 86 (24) | 70 (24) | 16 (24) |
| | Culture-confirmed on study sputum | 208 (57) | 175 (59) | 33 (50) |
| | Ultra-burden medium/high on study sputum | 106 (29) | 89 (30) | 17 (26) |

| | | | | |
|---|---|---|---|---|
| | Median Ct value (IQR) [§§] | 20.3 (17.9 – 26.3) | 20.3 (17.9 – 26.4) | 20.3 (18.4 – 26.2) |
| | Median time-to-culture-positivity (IQR) in days | 11.1 (7.1 – 17.3) | 10.9 (7.1 – 16.9) | 11.2 (7.3 – 20.2) |
| Radiological status | Median CAD4TB v6 score (IQR) | 78 (48 – 96) | 78 (49 – 97) | 70 (45 – 93) |
| | Cavitation | 59 (19) | 54 (20) | 5 (10) |
| | Advanced extent of disease** | 92 (41) | 79 (40) | 13 (45) |

PLHIV people living with HIV; ART antiretroviral treatment; IQR interquartile range; HDSS health and demographic surveillance system; WHO SS World Health Organization TB symptom screen; BMI body mass index

* VL unknown but on ART for > 6 months and CD4 count > 200 (n=10)

[§] Microbiological status based on samples collected on day of treatment initiation as part of study

[§§] Median of lowest Ct value for 4 *rpoB* probes given *M. tuberculosis* detected on at least one Ultra test

** Defined as per National Tuberculosis Association (NTA) classification (NTA-III)[8]

#### Table S2. Characteristics of child household contacts overall and by HIV and ART status of index PWTB

| | | All<br>N = 808<br>(col %) | HIV-negative<br>N = 271<br>(col %) | HIV+ on effective ART<br>N = 306<br>(col %) | HIV+ not on effective ART<br>N = 231<br>(col %) |
|---|---|---|---|---|---|
| Age (years) | Median (IQR) | 9.1 (5.9 – 11.9) | 8.9 (6.1 – 11.7) | 9.4 (5.9 – 12.1) | 9.0 (5.8 – 11.7) |
| Gender | Male | 420 (52) | 131 (48) | 157 (51) | 132 (57) |
| Geographical area in HDSS | East | 322 (40) | 98 (36) | 120 (39) | 104 (45) |
| | North | 237 (29) | 66 (24) | 110 (36) | 61 (26) |
| | West | 249 (31) | 107 (40) | 76 (25) | 66 (29) |
| Child HIV status | Negative* | 574 (71) | 193 (71) | 212 (69) | 109 (73) |
| | Positive | 12 (2) | 3 (1) | 3 (1) | 6 (3) |
| | Unknown | 222 (27) | 75 (28) | 91 (30) | 56 (24) |
| | *Declined HIV testing* | *307 (34)* | *104 (38)* | *126 (41)* | *77 (33)* |
| Child symptom screen positive§ | | 53 (7) | 27 (10) | 13 (4) | 13 (6) |
| Relationship to index | Child | 250 (31) | 61 (22) | 122 (40) | 67 (29) |
| | Sibling | 223 (27) | 98 (36) | 54 (18) | 71 (31) |
| | Grandchild | 151 (19) | 49 (18) | 71 (23) | 31 (13) |
| | Other family | 184 (23) | 63 (24) | 59 (19) | 62 (27) |
| Shared room with index PWTB (N = 704) | | 459 (65) | 146 (62) | 184 (70) | 129 (63) |
| QFT status | QFT-positive | 138 (17) | 60 (22) | 48 (16) | 30 (13) |
| | Indeterminate | 42 (5) | 20 (7) | 9 (3) | 13 (6) |
| | Insufficient sample | 11 (1) | 1 (<1) | 7 (2) | 3 (1) |

HDSS Africa Health Research Institute Health and Demographic surveillance system

* ELISA test or documented in Road to Health card or self-report

§ Child symptom screen includes cough (any duration), fever, night sweats, loss of appetite; plus age-specific symptoms—fatigue and weight loss (≥5 yrs), or decreased playfulness and failure to thrive (<5 yrs)

#### Table S3. Index PWTB-level factors associated with MTBC immunoreactivity

| | | Proportion QFT-positive N = 755 | Univariable association (adjusted for household clustering using GEE) |
|---|---|---|---|
| Index case demographics | | | |
| Age | 15 – 24 | 0.31(39/127) | 1 |
| | 25 – 34 | 0.20 (56/278) | 0.60 (0.31 – 1.16) |
| | 35 – 44 | 0.13 (22/165) | 0.37 (0.17 – 0.78) |
| | 45 - 54 | 0.07 (6/82) | 0.17 (0.06 – 0.44) |
| | 55 - 64 | 0.20 (10/50) | 0.49 (0.18 – 1.28) |
| | 65+ | 0.09 (5/53) | 0.20 (0.06 – 0.74) |
| Age continuous | Median (IQR) | 36 (29 – 47) | 0.97 (0.95 – 0.99) * |
| Sex | Male | 0.13 (47/361) | 1 |
| | Female | 0.23 (91/394) | 1.88 (1.17 – 3.02) |
| Education: grade 12 and above | No | 0.12 54/436 | 1 |
| | Yes | 0.26 84/319 | 2.46 (1.53 – 3.95) |
| Work status: unemployed | No | 0.22 (34/157) | 1 |
| | Yes | 0.17 (104/598) | 0.80 (0.48 – 1.33) |
| Different definitions of HIV status | | | |
| 1. HIV status at TB diagnosis: ART | HIV- | 0.24 (60/250) | 1 |
| | PLHIV on effective ART | 0.16 (48/290) | 0.51 (0.30 – 0.87) |
| | PLHIV not on effective ART | 0.14 (30/215) | 0.61 (0.32 – 1.15) |
| 2. HIV status at TB diagnosis: binary | HIV-negative | 0.24 (60/250) | 1 |
| | PLHIV | 0.16 (78/505) | 0.54 (0.33 – 0.88) |
| 3. HIV status at TB diagnosis: CD4 | HIV-negative | 0.24 (60/250) | 1 |
| (n = 748) | PLHIV CD4 ≥700 | 0.17 (20/121) | 0.47 (0.22 – 1.00) |
| | PLHIV CD4 500 - 699 | 0.08 (9/114) | 0.27 (0.11 – 0.69) |
| | PLHIV CD4 200 - 499 | 0.20 (28/141) | 0.72 (0.38 – 1.37) |
| | PLHIV CD4 < 200 | 0.16 (20/122) | 0.68 (0.35 – 1.31) |
| Symptom status | | | |
| Number of WHO TB symptoms | 0 | 0.14 (12/88) | 1 |
| | 1 | 0.11 (15/136) | 1.34 (0.45 – 4.00) |
| | 2 | 0.20 (32/160) | 2.46 (0.94 – 6.41) |
| | 3 | 0.19 (42/217) | 2.29 (0.87 – 6.01) |
| | 4 | 0.24 (37/154) | 2.55 (0.95 – 6.80) |

| | | | |
|---|---|---|---|
| Number of WHO TB symptoms | Median (IQR) | 2 (1 – 3) | 1.22 (1.02 – 1.47) ** |
| Different definitions of asymptomatic status | | | |
| 1. Symptom status: any symptom | At least one reported symptom | 0.19 (129/699) | 1 |
| | No reported symptoms | 0.16 (9/56) | 0.41 (0.12 – 1.39) |
| 2. Symptom status: WHO SS | At least one reported WHO symptom | 0.19 (126/667) | 1 |
| | Negative on WHO SS | 0.14 (12/88) | 0.45 (0.18 – 1.09) |
| 3. Symptom status: cough | Reported cough of any duration | 0.21 (111/529) | 1 |
| | Reported no cough | 0.12 (27/226) | 0.43 (0.25 – 0.74) |
| Clinical predictors | | | |
| Previous history of TB (N = 753) | No previous TB | 0.16 (92/564) | 1 |
| | History of previous TB | 0.24 (46/189) | 1.29 (0.80 – 2.10) |
| BMI | <18.5 | 0.22 (31/142) | 1 |
| | 18.5 – 24.9 | 0.21 (76/366) | 0.77 (0.42 – 1.42) |
| | 25.0 – 29.9 | 0.15 (20/136) | 0.51 (0.24 – 1.06) |
| | ≥30.0 | 0.10 (11/111) | 0.34 (0.12 – 0.93) |
| BMI continuous | Median (IQR) | 22 (19.5 – 26.4) | 0.95 (0.91 – 0.99) |
| Microbiological predictors | | | |
| Diagnostic (programmatic) sputum Ultra | Positive | 0.22 (125/560) | 1 |
| | Trace | 0.07 (13/195) | 0.27 (0.15 – 0.52) |
| Culture status (n = 751) | Culture-negative | 0.08 (26/316) | 1 |
| | Culture-positive | 0.26 (111/435) | 3.97 (2.31 – 6.81) |
| Days-to-culture-positivity (n = 235) | Median (IQR) | 11.2 (6.9 – 17.5) | 0.93 (0.87 – 0.99) |
| Xpert Ultra semi-quantitative output | Not detected | 0.08 (23/285) | 1 |
| | Trace | 0.10 (4/40) | 1.30 (0.35 – 4.77) |
| | Very low | 0.16 (16/101) | 2.03 (0.91 – 4.56) |
| | Low | 0.16 (19/116) | 2.38 (1.10 – 5.12) |
| | Medium | 0.30 (22/74) | 4.50 (1.80 – 11.25) |
| | High | 0.39 (54/139) | 7.41 (3.96 – 13.85) |
| Lowest Ct value of 4 *rpoB* probe (n = 430) [§] | Median (IQR) | 20.8 (17.9 – 27.3) | 0.89 (0.83 – 0.95) |
| Radiological predictors (n = 672) | | | |
| CAD4TB category | < 50 | 0.09 (17/186) | 1 |
| | 50 – 64 | 0.16 (14/88) | 1.50 (0.59 – 3.80) |

| | | | |
|---|---|---|---|
| | 65 – 79 | 0.16 (15/94) | 2.11 (0.82 – 5.46) |
| | 80 – 89 | 0.21 (18/86) | 2.47 (1.01 – 6.05) |
| | ≥90 | 0.27 (59/218) | 3.71 (1.83 – 7.50) |
| CAD4TB Score (continuous) | Median (IQR) | 75 (48 – 96) | 1.23 (1.09 – 1.38) *** |
| Cavitation (%) | Absent | 0.15 (83/539) | 1 |
| | Present | 0.30 (40/133) | 2.74 (1.59 – 4.73) |

MTBC *Mycobacterium tuberculosis*; QFT QuantiFERON Gold plus assay; GEE generalised estimating equations; IQR interquartile range; WHO SS World Health Organization TB symptom screen; BMI body mass index; Ct cycle threshold; CAD4TB output from computer aided detection of tuberculosis software

§ Median of lowest Ct value for 4 *rpoB* probes given *M. tuberculosis* detected on at least one Ultra test

* Assuming linear term for age

** Test for trend

*** Per 10-unit increments

#### Table S4. Child-level and household-level factors associated with MTBC immunoreactivity

| | | Proportion QFT-positive (n/N) N = 755 | Univariate association (adjusted for household clustering using GEE) |
|---|---|---|---|
| Child household contact | | | |
| Age in years | 2 – 4 | 0.13 (16/125) | 1 |
| | 5 – 9 | 0.14 (44/312) | 1.17 (0.72 – 1.90) |
| | 10 -14 | 0.25 (78/318) | 2.37 (1.40 – 4.01) |
| Gender | Male | 0.15 (59/385) | 1 |
| | Female | 0.21 (79/370) | 1.62 (1.17 – 2.25) |
| Relationship to index | Child | 0.21 (48/232) | 1 |
| | Sibling | 0.19 (39/208) | 1.08 (0.63 – 1.86) |
| | Grandchild | 0.14 (20/144) | 0.66 (0.35 – 1.26) |
| | Other family | 0.18 (31/171) | 0.75 (0.39 – 1.43) |
| Degree of exposure to index | Sharing room | 0.20 (87/432) | 1 |
| (n = 654) | Other | 0.17 (38/222) | 0.73 (0.44 – 1.20) |
| Household characteristics | | | |
| Geographical area (%) | North | 0.13 (29/227) | 1 |
| | West | 0.18 (43/234) | 1.29 (0.66 – 2.54) |
| | East | 0.22 (66/294) | 2.10 (1.21 – 3.66) |
| Household crowding (>2 persons / room) | No | 0.17 (81/485) | 1 |
| (n=743) | Yes | 0.19 (49/258) | 2.70 (0.84 – 8.67) |
| Socioeconomic status category[§] | Low | 0.14 (25/179) | 1 |
| | Middle | 0.16 (48/297) | 1.22 (0.64 – 2.33) |
| | High | 0.20 (111/665) | 1.64 (0.84 – 3.20) |

MTBC *Mycobacterium tuberculosis*; QFT QuantiFERON Gold plus assay; GEE generalised estimating equations

[§] Socioeconomic status category created using polychoric PCA

#### Table S5. Within-household MTBC transmission risk by ART status using GEE logistic regression with exchangeable correlation matrix and robust standard errors

| ART status (n = 467 contacts of 180 index PLHIV) | Number of index persons | Proportion child contacts QFT-positive (n/N) | Univariable odds ratio (OR) (95% CI) | Adjusted OR [a] (95% CI) | p-value |
|---|---|---|---|---|---|
| On effective ART | 103 | 0.147 (38/259) | Referent | Referent | 0.64 |
| Not on effective ART | 77 | 0.144 (30/208) | 1.31 (0.70 – 2.46) | 1.18 (0.59 – 2.34) | |

MTBC *Mycobacterium tuberculosis;* ART antiretroviral treatment; GEE generalised estimating equations; QFT QuantiFERON Gold plus assay; CI confidence interval

[a] Model includes *a priori* covariates (index age, index sex, geographical area, household socioeconomic status) based on directed acyclic graph

#### Table S6. Sensitivity analysis using TMLE: restricted to children aged < 11 years

| ART status (n = 301 contacts of 152 index PLHIV) | Number of index persons | Proportion child contacts QFT-positive (n/N) | Predicted probability of MTBC immunoreactivity [a] (95% CI) | Risk ratio[a] (95% CI) | p-value |
|---|---|---|---|---|---|
| On effective ART | 87 | 0.10 (16/166) | 0.089 (0.039; 0.140) | Referent | 0.35 |
| Not on effective ART | 65 | 0.11 (15/135) | 0.126 (0.066; 0.187) | 1.42 (0.68 – 2.95) | |

TMLE targeted minimum loss-based estimation; ART antiretroviral treatment; QFT QuantiFERON Gold plus assay; CI confidence interval

[a] Model includes *a priori* covariates (index age, index sex, geographical area, household socioeconomic status) based on directed acyclic graph

#### Table S7. Missing indicator method using TMLE

| ART status (n = 505 contacts of 194 index PLHIV) | Number of index persons | Proportion child contacts QFT-positive (n/N) | Predicted probability of MTBC immunoreactivity [a] (95% CI) | Risk ratio[a] (95% CI) | p-value |
|---|---|---|---|---|---|
| On effective ART | 113 | 0.166 (48/290) | 0.156 (0.100; 0.211) | Referent | 0.77 |
| Not on effective ART | 81 | 0.140 (30/215) | 0.145 (0.093;0.197) | 1.07 (0.66; 1,74) | |

TMLE targeted minimum loss-based estimation; ART antiretroviral treatment; QFT QuantiFERON Gold plus assay; CI confidence interval

[a] Model includes *a priori* covariates (index age, index sex, geographical area, household socioeconomic status) based on directed acyclic graph

#### Table S8. Characteristics of index participants by HIV and sex status

| | | Male | | Female | |
|---|---|---|---|---|---|
| | | PLHIV<br>N = 86<br>col % | HIV-negative<br>N = 61<br>col % | PLHIV<br>N = 108<br>col % | HIV-negative<br>N = 41<br>col % |
| Age (years) | Median (IQR) | 40.0 (33.2 – 50.8) | 35.0 (26.0 – 57.0) | 35.0 (30.0 – 46.0) | 27.0 (23.0 – 37.0) |
| Education | Grade 12 minimum | 29.1 | 39.3 | 47.2 | 63.4 |
| Employment status | Unemployed | 68.6 | 78.7 | 83.3 | 80.5 |
| Smoking status | Within last year | 41.9 | 32.8 | 3.7 | 0 |
| Geographical area in | East | 41.9 | 39.3 | 48.1 | 34.1 |
| HDSS | North | 31.4 | 32.8 | 26.9 | 26.8 |
| | West | 26.7 | 27.9 | 25.0 | 39.0 |
| Previous history of TB | | 37.2 | 8.2 | 30.6 | 14.6 |
| Definition of | No symptoms | 8.1 | 1.6 | 12.0 | 0 |
| asymptomatic TB | Negative on WHO SS | 10.5 | 4.9 | 19.4 | 4.9 |
| | No cough | 30.2 | 21.3 | 39.8 | 19.5 |
| BMI median (IQR) | | 20.8 (18.8 – 24.4) | 20.1 (17.9 – 22.5) | 24.6 (21.4 – 29.3) | 22.5 (20.4 – 26.4) |
| Microbiological status[§] | Diagnostic (programmatic) Ultra trace | 22.1 | 19.7 | 29.6 | 17.1 |
| | Ultra: not detected | 36.1 | 19.7 | 49.1 | 26.8 |
| | Ultra: medium/high | 29.1 | 47.5 | 15.7 | 43.9 |
| | Ultra: median Ct value (IQR)[§§] | 21.4 (18.0 – 26.6) | 18.9 (17.8 – 21.1) | 24.7 (18.0 – 28.2) | 18.6 (17.4 – 22.9) |
| | Culture-confirmed | 62.8 | 77.1 | 42.6 | 68.3 |
| | Median days-to-culture-positivity (IQR) | 12.6 (7.7 – 17.2) | 10.4 (6.6 – 14.3) | 12.8 (6.9 – 17.7) | 9.5 (6.6 – 12.2) |
| Radiological status | Median CAD4TB v6 score (IQR) | 79 (55 – 97) | 90 (79 – 98) | 53 (44 – 82) | 83 (49 – 99) |

| | | | | |
|---|---|---|---|---|
| Cavitation | 16.6 | 34.5 | 9.2 | 34.2 |
| Advanced extent of disease** | 34.5 | 58.2 | 24.1 | 55.3 |

PLHIV people living with HIV; ART antiretroviral treatment; IQR interquartile range; HDSS health and demographic surveillance system; WHO SS World Health Organization TB symptom screen; BMI body mass index

* VL unknown but on ART for > 6 months and CD4 count > 200 (n=10)

§ Microbiological status based on samples collected as part of study on day of treatment initiation other than programmatic (diagnostic) sample

§§ Median of lowest Ct value for 4 *rpoB* probes given *M. tuberculosis* detected on at least one Ultra test

** Defined as per National Tuberculosis Association (NTA) classification (NTA-III)[8]

#### Table S9. Household-attributable risk of child MTBC immunoreactivity: index versus matched control households (post-hoc TMLE analysis restricted to households registered in the AHRI HDSS)

| Household type | No. households | No. child contacts | QFT-positive, n/N (%) | Predicted probability of QFT-positivity (95% CI) [a] |
|---|---|---|---|---|
| Index households | 106 | 264 | 55/264 (0.21) | 0.20 (0.15 – 0.24) |
| Control households | 111 | 189 | 12/189 (0.06) | 0.08 (0.06 – 0.10) |
| Risk difference, index − control (proportion, 95% CI) [a] | | | | 0.11 (0.08 – 0.15) |
| Household-attributable fraction, % (95% CI) [b] | | | | 58.7 (47.4 – 67.4) |

MTBC = *Mycobacterium tuberculosis* complex; QFT = QuantiFERON-TB Gold Plus; TMLE = targeted minimum loss-based estimation; CI = confidence interval.

[a] TMLE predicted QFT-positivity and the risk difference were estimated by TMLE with an ensemble machine-learning library, accounting for household-level clustering and adjusting for child age, child sex, household crowding (persons per sleeping room) and geographical area. The risk difference is the difference in expected proportion of immunoreactivity had all children resided in index versus control households.

[b] Household-attributable fraction = percentage of MTBC immunoreactivity among children in index households attributable to co-residence with a person with bacteriologically-confirmed pulmonary tuberculosis, over and above the community-acquired background captured by control households

### References

1. Banaei N, Gaur RL, Pai M. Interferon Gamma Release Assays for Latent Tuberculosis: What Are the Sources of Variability? J Clin Microbiol 2016;54(4):845–50.

2. Nemes E, Rozot V, Geldenhuys H, et al. Optimization and Interpretation of Serial QuantiFERON Testing to Measure Acquisition of M. tuberculosis Infection. Am J Respir Crit Care Med 2017;

3. Dang LE, Gruber S, Lee H, et al. A causal roadmap for generating high-quality real-world evidence. J Clin Transl Sci 7(1):e212.

4. Hernan MA, Robins JM. Causal Inference: What If [Internet]. Chapman & Hall/CRC; 2024. Available from: https://miguelhernan.org/whatifbook

5. Martel P, Mbofana F, Cousens S. The polychoric dual-component wealth index as an alternative to the DHS index: Addressing the urban bias. J Glob Health 11:04003.

6. Myburgh H, Peters RPH, Hurter T, Grobbelaar CJ, Hoddinott G. Transition to an in-facility electronic Tuberculosis register: Lessons from a South African pilot project. South Afr J HIV Med 2020;21(1):1025.

7. WorldPop (www.worldpop.org - School of Geography and Environmental Science, University of Southampton; Department of Geography and Geosciences, University of Louisville; Departement de Geographie, Universite de Namur) and Center for International Earth Science Information Network (CIESIN), Columbia University (2018). Global High Resolution Population Denominators Project - Funded by The Bill and Melinda Gates Foundation (OPP1134076). https://dx.doi.org/10.5258/SOTON/WP00645 [Internet]. Available from: www.worldpop.org

8. NTA. Diagnostic Standards and Classification of Tuberculosis [Internet]. National Tuberculosis Association; 1940. Available from: https://catalog.hathitrust.org/Record/001562842
